# Practical Challenges for Commercial Enterprises in the Ethics Review Process for Digital Health Research: Document Analysis and Interview Study

**DOI:** 10.1101/2024.01.28.24301885

**Authors:** Katherine Yang, Henry W W Potts

**Affiliations:** UCL Institute of Health Informatics, University College London

**Keywords:** Ethical review, Ethics committees, Informatics, Commerce, Public-private sector partnerships, Digital health

## Abstract

**Introduction:** The rapid evolution of digital health interventions has created challenges in navigating the ethics approval process for commercial enterprises. Recognising the need for processes that balance ethical considerations with the specifics of digital health research, this study aimed to describe what happens when enterprises seek ethical review in the UK and propose strategies for a smoother process.

**Methods:** Inductive thematic analysis was conducted on thirty-two ethics review documents (29 to an NHS Research Ethics Committee, 3 to an ethics committee at a higher education institution) submitted by digital health developers with commercial sponsors and ten semi-structured interviews with digital health enterprise representatives.

**Results:** Ethics committees raised an average of 4.3 action points per submission. We identified five broad themes around committees’ concerns: ethical commitments in care; study design; digital health research peculiarities; data governance; document quality and completeness. Interviewees reported a range of experiences. Here, we identified six broad themes: submission and protocol revisions; the dynamic between parties; application time and procedures; acumen and practicality in digital health; support and guidance from RECs; enterprise expertise and resources.

**Conclusion:** We suggest strategies for applicants to achieve a favourable decision, such as evidence-based study designs and participant support for better inclusion and equity, and identified specific pitfalls to avoid, such as lack of justification for data governance procedures. We recommend that UK research ethics committees provide adapted guidance and foster collaboration through open communication and mutual understanding, to facilitate a smoother approval process in digital health research.

## Introduction

Digital health encompasses the use of a wide range of technology, including mobile apps, wearables, and telemedicine, to enhance health outcomes and services ^1, 2^. Recent years have witnessed accelerated developments in digital health technology, propelled by pandemic initiatives ^3^. While digital health interventions have vast potential, there is concern that they are infrequently and poorly evaluated ^4^. The UK has made strides to enhance the evaluation of digital health technology ^5, 6^, exemplified by the NICE Evidence standards framework for digital health technologies ^7, 8^.

Research ethics committees (RECs, also known as Institutional Review Boards in the US) scrutinise the ethical aspects of research proposals and provide a formal opinion. Since the 1960s, RECs have safeguarded human subjects’ rights ^9,10^, adhering to codes of ethics from professional associations and educational institutes to guide standards ^11, 12^.

Research ethics is not governed by a single legislation in the UK. However, obtaining ethical approval before commencing on a research study from entities like UK Research Ethics Service (RES) or higher education institutions (HEIs) is essential for journal publication and further implementation of results ^13, 14^. The NHS Health Research Authority (HRA) in England and equivalents in the devolved administrations collaborate on the Research Ethics Service (RES), overseeing multiple RECs across the UK, providing reviews for research involving NHS patients, staff, or facilities. This national system represents more centralisation than, for example, the North American context. Applications to RES RECs are made via a web portal, the Integrated Research Application System (IRAS) ^15^. Applications are assigned to a REC and reviewed during a meeting, where concerns are addressed, questions posed, and opinions given. A favourable opinion can be given, but the REC will often ask for amendments or further review ^16^.

HEIs maintain their own RECs to review research by affiliates, when it falls outside the HRA’s scope. The specifics of the process and REC structure vary between institutions ^17^. Additionally, major corporations, like pharmaceutical firms, often maintain internal governance boards.

While RECs traditionally focus on protecting participants and ensuring societal benefits ^10^, modern healthcare’s evolution and the advent of digital health technology introduced new ethical complexities, such as juggling privacy with the need for public health surveillance, data ownership concerns, consent validity, and disparities from algorithmic biases ^18, 19^. Balancing stakeholders’ interests at the crossroads of participants, researchers, and society ^20^, RECs are often vulnerable to being received as disrespectful, bureaucratic or obstructive ^21^. Past studies suggest that perceived unfairness might tempt researchers towards misconduct to “level the playing field”, leading to negative consequences ^22^. From the REC’s perspective, researchers might seem to be merely “checking boxes” for regulatory compliance rather than genuinely addressing ethical implications ^23^. A collaborative, respectful partnership between researchers and RECs benefits all parties involved, minimising unrealised research potential and promoting research quality and integrity ^22^.

Recent times have marked a surge in industry-funded digital health trials ^24, 25^. Relevant and timely evaluation is crucial to keep pace with technological advancements. This study drew inspiration from unpublished work asking digital health developers their views on evaluation, which pinpointed ethics applications as a significant challenge (Paulina Bondaronek, pers. comm.). Enterprises often cite issues like data ownership, fair access, consent during data collection, and relationships with RECs as obstacles to meeting ethical standards.

Over the past decade, research ethics guidelines have adapted to digital innovations, offering more up-to-date tools and guidance ^26^. Recent global guidelines underscore the values and principles of ethics in digital health research, highlighting the need to stay updated on ever-evolving study designs ^27–31^. However, a standardised framework tailored to address ethical concerns in digital health has yet to be developed ^28^. Researchers have critiqued the disconnect between current regulations and the actualities of digital health research, where blurring boundaries, such as between commercial and non-commercial work, cause uncertainties ^32, 33^. Even REC members report feeling uncertain about digital health research ethics ^34^.

The ethical review process is often seen as confusing due to the absence of standardised procedures. While the majority of healthcare researchers report being confused by ethical ambiguities, only a few regularly consult RECs for guidance ^35^. Criticisms of RECs are common in research literature ^36–38^. While there are none in the context of digital health, numerous studies explored the ideal REC role, reinforcing the importance of ethical approval in research ^21, 22^. Previous studies have often limited their scope to a subset of digital health or a specific ethical challenge ^18, 39^, leaving a gap in a comprehensive examination of the ethical approval process in digital health to tackle practical challenges.

The aim of this investigation was to identify and understand challenges encountered by digital health enterprises in the UK during the research ethics approval process. By analysing formal documentation outlining REC responses to submitted applications, the study determines common barriers. Interviews with enterprise representatives provide an understanding of their direct experiences and challenges faced.

## Methods

The study had two parts in parallel: document analysis and interviews with representatives from enterprises. Participants could engage in either or both approaches. The study received ethical approval from the UCL Institute of Health Informatics REC (8-IHILREC).

### Ethics Application Documents

Individuals submit an ethics application form via IRAS to RES RECs. These are discussed in the REC’s meetings, which are minuted. This can lead to a favourable opinion (the study can proceed), a favourable opinion with conditions (the study can proceed after certain conditions are met), a provisional opinion (where the applicant is asked to make revisions for subsequent review), or an unfavourable opinion. A response to the applicant with action points is produced. The opinion and action points to be communicated are detailed in the meeting minutes.

Applications to RECs can be submitted as being of lower risk for a more streamlined application process, called Proportionate Review. HEI RECs may have similar arrangements.

Digital health research ethics applications with a commercial sponsor reviewed by a RES REC in the UK between May 2020 and May 2023 were identified with the following keywords: wearable, informatics, digital health, mobile health, information technology, telehealth, telemedicine, personalised medicine. Drug trials were excluded. The keywords were refined in discussion with the HRA to ensure a practical number of applications could be identified. The search produced 33 applications.

At the suggestion of the HRA, a Freedom of Information (FOI) request for completed application forms and ethics committee meeting minutes (summarising discussions between the REC members and applicants), provisional requirements, decisions and recommendations was made to the HRA and extended to the identified applicants. Of the identified applicants, 27 did not decline the FOI request and their anonymised documents were provided.

Another five sets of documents, including ethics application forms and responses from RECs, were shared by interview participants (details below). Two of these were submitted to the HRA and three to an HEI REC in the UK. One document concerned a project involving one of the authors (HP). These documents were anonymised by KY.

Combined, a total of 32 ethics applications submitted to various RECs across the UK and the committees’ responses were collected and analysed through thematic analysis to identify common challenges in ethical approval.

### Applicant Interviews

Commercial enterprises were invited to semi-structured interviews if they met inclusion criteria of being registered and operating in the UK; conducting research in digital health; requiring ethics approval from an external REC instead of through an internal process; and having submitted a research ethics application in the past 5 years.

We first approached digital health enterprises with whom a previous collaborative relationship existed. Interview participants were also recruited through advertisements posted by our personal accounts on LinkedIn and Twitter (although no responses came via Twitter). A snowball technique was used whereby identified participants could refer their peers. An invitation email outlining research objectives and procedure was sent to those who expressed interest, followed by the participant information sheet (PIS) and consent form if they were willing to participate. This produced six participants.

In the process of obtaining ethics application documents through the FOI request via the HRA, several further participants were identified. These individuals agreed not only to share their application documents but also to participate in interviews. Four applicants corresponding to six sets of documents thus also participated in interviews.

Overall, ten interviews were conducted. Before the interviews, consent was obtained from each participant, ensuring they were fully aware of the research objectives, the nature of their participation, the confidentiality of their information, and their right to withdraw at any point without consequences.

The interviews lasted 30-60 minutes and took place on MS Teams or Zoom. Transcripts were produced using the platforms’ own functions, then verified against the recordings for accuracy. Identifiers in the transcripts were removed. The semi-structured interview topic guide, formulated based on past research and current study objectives, is in Appendix II.

### Document and Transcript Analysis

Thematic analysis ^40, 41^ was conducted on the REC meeting minutes or decision letters, outlining the REC’s response to the application. Other information, including the study patient population, whether the REC was designated for medical device studies, whether the application was submitted for a low-risk proportionate review, the decision (favourable, favourable with additional conditions, provisional, or unfavourable opinion), and the actions required by the ethics committee, was also noted. The IRAS application forms were not coded, but were reviewed as needed to understand the research projects and committee comments. These forms served as a reference to how and if applicants addressed certain topics and to recognise effective practices that prevented common issues. A second, but complementary thematic analysis was conducted on the interview transcripts.

Coding and subsequent analysis were done in NVivo 14 Plus. Initial codes were generated after reviewing the documents or interview transcripts, individually and separately, through a data-grounded inductive approach to produce rich, detailed themes. A passage could be coded to multiple codes. Related codes were clustered into potential themes, after which the themes were iteratively reviewed, edited, defined and named. The themes generated from the documents and the interview transcripts were compared and contrasted to identify overlaps.

Cross-coding with a secondary coder was conducted for randomly selected documents to ensure consistency and validity. The primary coder’s codes and themes were provided as reference. The secondary coder had the freedom of including additional codes and organising the codes into subthemes as seen fit. The coders discussed conflicts or mismatches between the coding results, and all disagreements were resolved.

## Results

### Document Analysis Findings

#### Application Characteristics

About half of the ethics applications (17/32) received a decision with provisional opinions and four (13%) received favourable opinion. None received an unfavourable opinion. Eighty percent (25/32) of the studies involved non-healthy adult participants; five involved children/adolescents; two had healthy participants.

The outcome of the discussions in the review meetings were categorised as: satisfactory responses, optional recommendations, or actions points needing amendments. Over all the applications, there was an average of 4.3 actions point (range 0-16). The applications given a provisional opinion had 4.9 action points on average compared to 1.8 for those receiving a favourable opinion.

**Table 1.** Summary of key attributes of the ethics review meeting minutes.

|  | *Favourable Opinion*<br>*(n=4)* | *Favourable Opinion*<br>*with Conditions*<br>*(n=11)* | *Provisional Opinion*<br>*(n=17)* |
| --- | --- | --- | --- |
| **REC Expertise: Medical Device Studies** |  |  |  |
| *Designated, HRA REC* | 1 | 2 | 3 |
| *Non-Designated, HRA*<br>*REC* | 3 | 9 | 11 |
| *Non-Designated,*<br>*Non-HRA REC* | 0 | 0 | 3 |
| **Participant Population** |  |  |  |
| *Healthy Adult*<br>*Subjects* | 0 | 0 | 2 |
| *Non-Healthy Adult*<br>*Subjects* | 3 | 9 | 13 |
| *Children and/or*<br>*Adolescents* | 1 | 2 | 2 |
| **Number of Action Points** |  |  |  |
| *Mean* | 1.8 | 4.5 | 4.9 |
| **Number of Proportionate Review** |  |  |  |
| *Count* | 1 | 1 | 1 |

### Common Concerns and Barriers

We identified five main themes and 16 subthemes (Figure 1), detailed below. Example quotations are highlighted in tables.

**Figure 1.**
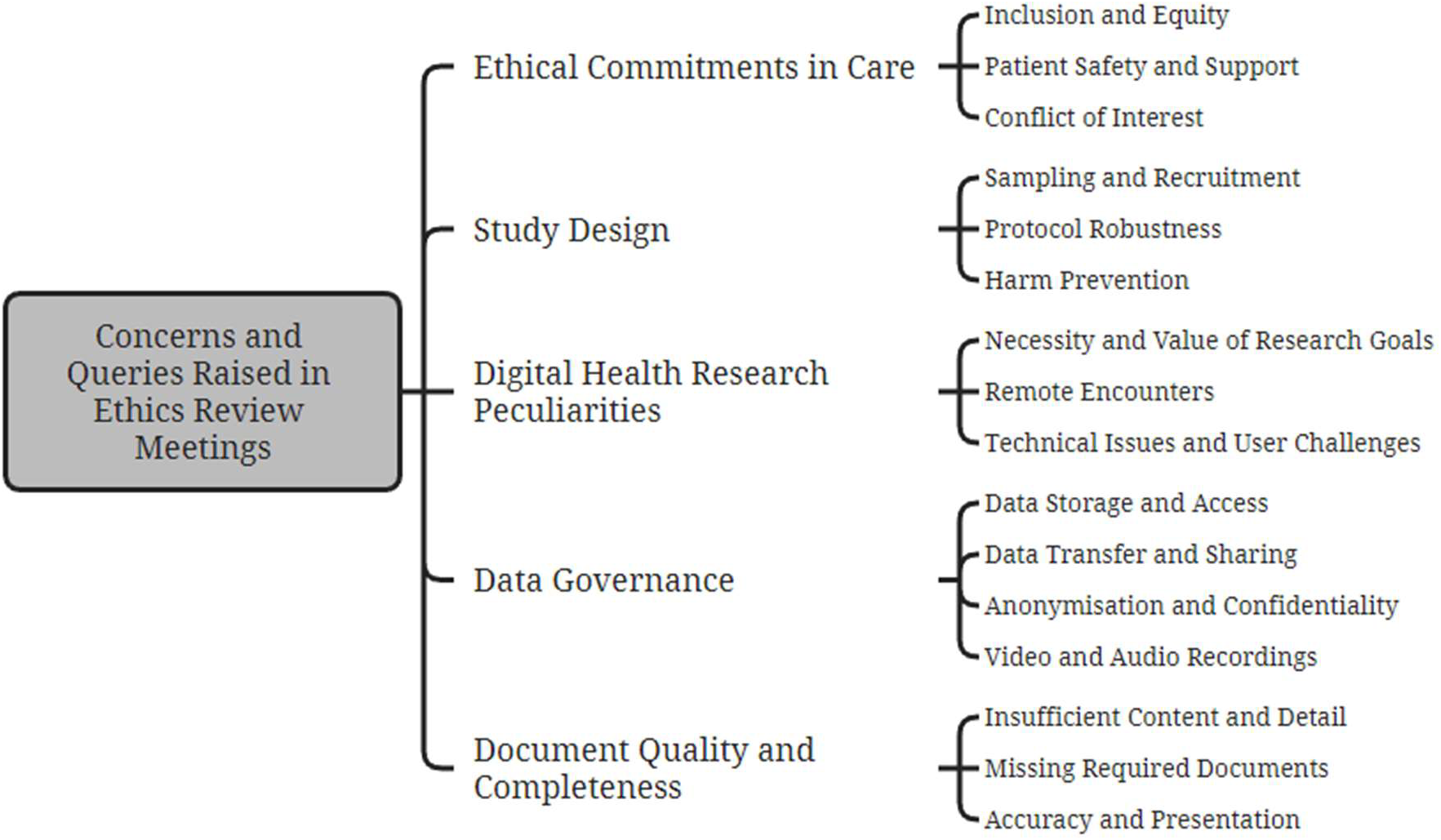
Themes and subthemes identified from ethics committee meeting minutes, detailing the concerns and queries raised.

**Figure 2.**
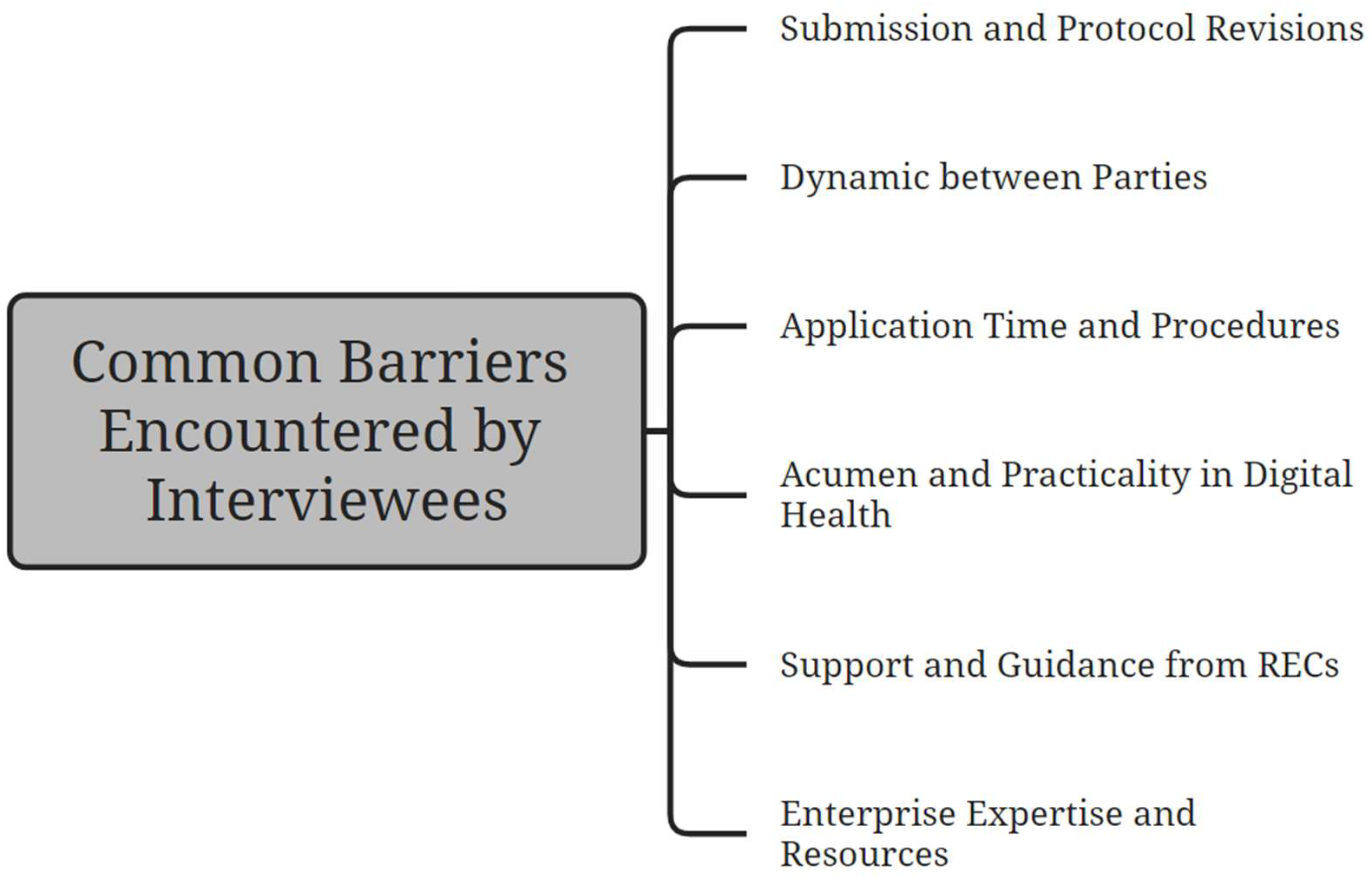
The themes identified from interviewee discussions of barriers and challenges encountered during the ethics application process.

### Theme 1: Ethical Commitments in Care

**Table 2.** Summary of discussion topics and outcomes under the theme ‘Ethical Commitments in Care’.

| Subtheme | number of action points | number of satisfactory responses | number of applications | Codes |
| --- | --- | --- | --- | --- |
| Inclusion and Equity | 20 | 18 | 18 | Access to delivery platform<br>Access to physical device<br>Compensation and incentives<br>Cost burdens<br>Existing constraints<br>Physical and cultural demographics attributes<br>Language and cultural barriers<br>Special patient groups |
|  |  |  |  | Restriction due to relationships |
| Patient Safety and Support | 38 | 24 | 22 | Public and patient involvement<br>Workload and time burdens<br>Technological burdens<br>Pressure to participate<br>Safeguarding special patient groups and children<br>Safeguarding of others<br>Instruction on technology and other study procedures |
| Conflict of Interest | 16 | 0 | 7 | Transparency in financial involvement<br>Relationship with sponsor<br>Clarity on research beneficiary<br>Role of the researcher and bias |

#### Inclusion and Equity

The REC inquired about the availability of smartphones, tablets, internet access, video conferencing tools, and travel cost reimbursements, requesting provisions where needed and flagging omissions as action points to ensure equity. Withdrawing access to the service or device post-study was also identified as a risk for impeding participation and impacting subsequent care. Compensations that were inequitably low or varied between new and existing users were flagged for amendments.

Concerns arose when software was limited to specific operating systems, necessitating justifications and future rollout plans. A commercial health app’s pre-existing license agreement placed undue demands on participants, leading to a modification request.

For studies involving physical devices or software capturing physical features, the RECs inquired on the device’s adaptability to different body shapes and physical features. RECs emphasised the importance of inclusive questionnaire options beyond binary gender choices and sought justifications for exclusions based on BMI criteria. While including a broad spectrum of body types, sexes, and ages was highlighted, particularly in smaller sample sizes, these concerns generally did not lead to extensive amendments, as applicants often provided sufficient rationales for their design and participant selection choices.

Translations or interpreters were often sought for non-English speakers. Data collection was advised to be inclusive, accommodating various measurement systems and international qualifications. Justification was requested for the inclusion or exclusion of special patient populations like impaired and pregnant patients. RECs encouraged the applicants to be “as inclusive as possible in their recruitment” when needed. Discussions on these complex ethical issues are lengthier than others, but were of a supportive and constructive nature.

**Table 3.** Example quotations from meeting minutes on the ‘Inclusion and Equity’ subtheme, organised by codes.

| Code | Example Quotation |
| --- | --- |
| Access to delivery platform | "Clarifications were requested in relation to the choice to deliver this study solely through Apple products, and if the study team has any plans to develop this for other devices (Android) in the future." |
| Access to physical device | "The Committee highlighted the limitation of the digital divide, where some participants may not have access to technology to participate in the study. The applicant said they would look to create funding to provide devices. The Committee suggested approaching charities who provide technology support." |
| Cost burdens | "The Committee asked if travel expenses would be reimbursed. The applicant said they would minimise travel requirements by screening in the participant's usual primary care setting, so they would likely live nearby. The Committee requested that travel expenses be reimbursed as required." |
| Compensation and incentives | "30min = £6 per hour – this rate would need justification, ideally with evidence for why this is an appropriate amount, especially when existing users are not benefiting in the same way." |
| Existing constraints | "Please clarify that the participant is not expected to indemnify the sponsor, as the [app] end user license agreement and privacy policy implies the participant will be responsible for this, and cannot participate if they do not sign to agree." |
| Restriction due to relationships | "Please justify exclusion criterion 6 as this could disadvantage a potential participant who lives with, and is related to, a study staff member: The subject or a close relative of the subject is the investigator or a subinvestigator, research assistant, pharmacist, study coordinator, or other staff directly involved with the conduct of the study at that site." |
| Physical and cultural demographics attributes | “The Committee asked [for] more information on the inclusion/exclusion criteria, for example, would certain ethnic groups be included and what about people who had plastic surgery. Specifically, the Committee asked [...] why males were to be excluded and participants over a certain Body Mass Index (BMI). The Chief Investigator explained that males were partly excluded as it was established that females used more facial expressions than males. The researchers also highlighted that if men were included then they would require the glasses to be in various different sizes. The applicants explained that they would be excluding those who may have neurological or cognitive difficulties, as well as people who had surgery which affected facial expressions. Regarding the exclusion of participants over a certain BMI, the Chief Investigator explained that they wanted to recruit participants whose weight would not affect their movement. The Committee accepted this response.” |
| Language and cultural barriers | “Please review the parent/carer initial questionnaire, question 18 (education/qualification). Please make this question easier to answer for all participants who may not have been educated in the UK and ensure that this question takes into considerations qualifications gained from around the world and not just the UK.” |
| Special patient groups | “The members noted that in young adults with this condition there may be cognitive impairment [with] communication... dexterity problems preventing use of communication tools such as writing, hence very likely that some potential participants would be unable to give their own informed consent [...] The Committee agreed the research could not be carried out as effectively if it was confined to participants able to give consent [...] [as that] would exclude a significant group of potential participants.” |

#### Participant Safety and Support

About one fifth of the issues discussed surrounded public and patient involvement (PPI) in designing study protocols and developing intelligible materials. Most applicants had involved the public, while those who did not received either recommendations or action requirements to do so, depending on the suitability of the submitted information sheets and consent forms. Proactive PPI also alleviated committee concerns surrounding the burden of participation or possible frustration from using the technology.

Other amendments to the recruitment protocol were requested to reduce patient burden or lessen the pressure on potential participants, including patients and staff, to ensure voluntary participation. Control groups and those interacting with participants also needed safeguarding. Whenever discussions about delivering questions of a sensitive nature or to sensitive participant populations arose, all applicants addressed them by presenting well-considered arrangements and justifications.

Over a third of the applications faced queries regarding technology usage instructions. While some lacked clarity or were not user friendly, others, despite having clear guidelines in the application, had omitted them from participant information sheets. Such oversights consistently led to amendment requests. Depending on complexity, some studies needed face-to-face guidance on app usage and exercises, with helplines suggested for login troubles. High-risk devices also required laboratory tests and safety checks before approval.

**Table 4.** Example quotations from meeting minutes on the ‘Participant Safety and Support’ subtheme, organised by codes.

| Code | Example Quote |
| --- | --- |
| Public and patient involvement | "The Committee asked the applicant to confirm if they had received any PPI involvement. The applicant confirmed they had not received PPI involvement with the [PIS]. The Committee commented that PPI involvement in patient-facing documentation was keenly encouraged." |
| Workload and time burdens | "There are many questionnaires to complete... perhaps allow participants to complete the study in stages." |
| Technological burdens | "The Committee queried whether the researchers had considered whether the technology may frustrate and overburden participants who are unfamiliar with the technology.<br>"The researchers explained that they have previously conducted a good amount of research using this technology, with this specific software being used in their previous study that looked at software engagement. In their experience the use of this technology had not created a great burden or frustration, and in circumstances where participants are not using the software then the researcher would contact the participant to ask if they were having any issues." |
| Pressure to participate | "The Committee agreed that people should be free to choose to be screened for a medical condition, however this choice is taken from them when they would receive a letter stating they were at high risk of AF. The Committee added knowing this information could make it difficult for people to decline to take part in the study... The Committee asked if patients could highlight to their GP practice that they did not wish to be involved in research." |
| Safeguarding special patient groups and children | “The Committee asked the applicants if any feedback was taken from any patient groups with regards to the easy read version of the participant leaflet (PIS). The applicant explained that the easy read PIS was written in partnership with a group...who specialise in patients with autism and disabilities. The Committee accepted the response.” |
| Safeguarding of others | “Consider providing participants with a small card to hand out and warn people that any interaction will be recorded.” |
| Instruction on technology and other study procedures | “[The] Committee agreed that a separate information sheet for each specific device would need to be created that included instructions on how to use, charge, wear and care for the device... Each participant would be given the specific one that related to the device they had been given as part of the study.” |

#### Conflict of Interest

Although only affecting six out of the total 32 applications, nearly all issues surrounding conflicts of interest led to action points and unresolved queries. Applicants failed to include details in info sheets or adverts, including past and current affiliations with the sponsor, irrespective of their relevance to the study, and a clear explanation of the sponsor’s identity. One review requested confirmation that analysts were sponsor employees. The committee also requested that a researcher who is financially affiliated with the sponsor not be involved in consenting patients.

When submitted to a HEI REC, there was heightened emphasis on delineating relationships and dynamics between researchers, listed organizations, study locations, and the approving institute. The REC also emphasised incorporation of institutional affiliation on participant-facing materials.

**Table 5.** Example quotations from meeting minutes on the ‘Conflict of Interest’ subtheme, organised by codes.

| Code | Example Quote |
| --- | --- |
| Transparency in financial involvement | “The Information sheet should include detail on the personal and research institutional financial support received by the Chief Investigator from [the sponsor], for full transparency with participants.” |
| Relationship with sponsor | “Providing the raw data to [the sponsoring enterprise] — and not just the research results — makes this feel like consultancy. A possible solution: provide the sponsor with early access to the research results (prior to publication), but not the raw data itself.” |
| Clarity on research beneficiary | “The Information Sheet should more clearly set out how the research findings will be used and by who. It seems as though the primary beneficiary will be the company..., who plan on using the |
|  | findings to develop software for sale? How will Prostate Cancer UK use the findings?" |
| Role of the researcher and bias | "A4 implies that the main researcher is external to [the HEI]. Similarly, there ought to be some consideration that – at least as implied by B1 – the researcher appears to be testing the success of their own app." |

### Theme 2: Study Design

**Table 6.** Summary of discussion topics and outcomes under the theme ‘Study Design’.

| Subtheme | number of action points | number of satisfactory responses | number of applications | Codes |
| --- | --- | --- | --- | --- |
| Sampling and Recruitment | 27 | 14 | 20 | Inclusion and exclusion<br>Participant recruitment<br>Remote consent<br>Time given to consider consent<br>Sampling<br>Participant definition |
| Protocol Robustness | 12 | 8 | 14 | Interview design<br>Justification and rationale<br>Broader protocol adequacy<br>Compliance with care standards<br>Feedback and results distribution<br>External matters |
| Harm Prevention | 17 | 13 | 17 | Staff and clinician training<br>Unforeseen events<br>Non-adherence or withdraw<br>GP or specialist contact<br>Safety concerns<br>Participant and co-habitant contraindications |

#### Sampling and Recruitment

Issues arose with inconsistent and contradicting recruitment methods, such as recruiting from A&E for a study aimed at healthy volunteers, which would then call for a set of medical exclusion criteria due to their acute illnesses. There were gaps in the information in applications about age range, withdrawals, and justifications for excluding certain groups. Applicants had to provide additional consent forms and information sheets for carers, family, and non-team research staff, having initially overlooked their inclusion as participants. RECs also questioned the qualifications of clinicians assessing or recruiting patients, and scrutinized how well exclusion criteria were upheld when participants self-engaged. When researchers excluded patient groups due to their limited relevance to the study or its preliminary nature (not for strict CE marking), the REC found these reasons acceptable.

Many applications lacked adequate protocol descriptions for sampling and recruitment, with verbal clarifications during the meetings frequently leading to action points. Common areas needing detail included the initial approach strategy, method of invitation and consent, and meeting formats (in person or virtual). Justifications were often required, especially regarding sample size determinations. Some applicants justified approaching more participants by referencing past trials and recruitment success rates.

**Table 7.** Example quotations from meeting minutes on the ‘Sampling and Recruitment’ subtheme, organised by codes.

| Code | Example Quote |
| --- | --- |
| Inclusion and exclusion | “The Committee queried if people in the depressed group that were already on [anti-depressants] would be included in the study. The applicant confirmed individuals on anti-depressants could still take part as the researchers did not want to exclude these individuals and felt it was unethical to ask for people to stop their medication. The Committee queried whether the researchers would collect data on what medications were being used by participants in order to account for this during their analysis. The applicant confirmed that if the participant disclosed this themselves then this would be recorded, however the researchers would not be collecting this information intentionally. The Committee recommend the researchers consider whether it would be useful to collect data on any anti-depressant medication being used by participants and amend the application accordingly if the research team plans to collect this” |
| Participant recruitment | “The Committee were unsure how the Applicant intended to recruit as the application was inconsistent. The IRAS form stated recruitment was via an advertising poster but also stated it would be from A&E admissions. In further deliberation the Committee agreed that if recruitment was via A&E admission a medical perspective inclusion and exclusion criteria would be required. Furthermore, A&E recruitment also brought into question the study’s recruitment of healthy volunteers.” |
| Remote consent | “The Committee asked how consenting would be managed in light of any Covid restrictions. Confirmed that consenting could be done remotely via the online platform if necessary.” |
| Time given to consider consent | “Members queried the rationale for participants... only having one hour to decide on whether to participate.<br>“[The applicant] stated that 48 hours could be given to participants but as the potential participants were already in the A&E Department, [he/she] did not want to cause an inconvenience. [He/she] stated that the PPI Group had looked at this procedure and agreed that it was acceptable.<br>“Members stated that consideration must be given to allowing participants a minimum of 48 hours to decide on whether to participate in the research study.” |
| Sampling<br>Participant recruitment | “... the response... mentions that 10 participants will be asked to... participate... and this would include at least 10% of those who chose to drop out. The Committee noted that some of the questions may not be relevant for people who decide to drop out and it is also not clear if dropping out would mean withdrawal of consent in which case it may not be appropriate to re-approach them.<br>“[The applicant] explained that they will be very sensitive when approaching these patients, especially as these will be autistic patients. The research team will make sure the patients are comfortable about giving consent. He explained that one of the researchers, who is an autistic person, fully understands the sensitivity of the issues and will be involved in this process. He further explained that the 10% participants mentioned in the above statement will include those who will be willing to participate but had to drop out for some other reasons. This is only to understand the reasons for dropping out as it could be very important for research. The Committee accepted the response.” |
| Participant definition | “The Committee acknowledged that individuals providing care to the patients (i.e. carers) should also be considered as participants since they will be asked to complete the burden questionnaire. The members, therefore, requested a separate Participant Information Sheet and Informed Consent Form is created for this participant-group.” |

#### Protocol Robustness

The number of inquiries concerning the broader adequacy of the protocol was smaller than other subthemes, but often required intricate meeting discussions, demanding greater effort to address. Topics requiring justification for the protocol displayed a diverse range in both topic and nature, overlapping with subthemes mentioned previously. They spanned areas including the necessity for medical testing, potential technological burdens faced by participants, rationale for specific data collection, presumptions regarding positive trial outcomes, the design of interviews, feedback and results distribution, adherence to established care standards, and much more. Some issues even led to entire sections of the study protocol being removed. These multifaceted issues underscored the need for comprehensive reasoning and clarity in protocol development. The outcome of any inquiry—whether action was required or a response deemed satisfactory—was determined by the strength of the supporting rationale and the complexity of the presented issues.

**Table 8.** Example quotations from meeting minutes on the ‘Protocol Robustness’ subtheme, organised by codes.

| Code | Example Quote |
| --- | --- |
| Interview design | "The Committee noted that the questions in the interview schedule for the carers were quite prescriptive and specific about severity and frequency rather than being open ended, as expected in a qualitative study... The applicant explained that they have tried to use evidence-based approach in designing the discussion guide for the carer interviews... by reviewing the literature on care of burden on rare disease and ask the relevant questions based on that. [He/she] added that not much work has been done in this field and this is why they are using the existing evidence to guide them. The Committee...agreed..., however the follow-up questions are very specific...to level and frequency and could be made more general and exploratory. The applicant... agreed to revise the follow up questions by possibly reducing the element of quantification." |
| Justification and rationale | "[The PIS] mentioned TB testing..., but it was not clear why this was necessary as there was no other details in the IRAS form or Protocol... Justify the use of TB testing and other screening tests (e.g. LFT's, HIV, Hepatitis B) in this study." |
| Feedback and results distribution | "The Committee asked if the applicant would feedback to the participant whose test results determined that they were depressed but were not clinically diagnosed as depressed. The applicant responded that they would never feedback to the patients as they were not doctors. They would also not call it depression. If the participant needed help, they would be referred to their GP. The Committee was satisfied with this response." |
| Compliance with care standards | "The Committee asked to clarify who will make the decision... to continuation of the treatment for the patients[...] [The applicant] explained that[...] as a standard... it would be unusual if they were continuing with the treatment. However, this will have nothing to do with the study and the study procedures will not interfere with the clinical practice in any way as the decisions will be made by the treating clinicians. The Committee accepted the response." |
| Broader protocol adequacy<br>External matters | "The Committee questioned whether it would be appropriate to invite participants in a COVID19 human challenge study into an additional piece of research[...] The Committee were familiar with the complexity and ethical issues raised by conducting a human challenge study on a new virus that had become a global pandemic and was of the opinion that recruitment of this group would not be appropriate. Please therefore include this as an exclusion in the study protocol." |
|  | “Please note however that the REC would be willing to review a substantial amendment to include this group at a later date (if this was required in order to answer the research question), provided that all additional ethical issues (specifically related to inclusion of this group) have been considered and reasonably justified.” |

#### Harm Prevention

Most applications that encountered harm prevention issues (16/17) faced barriers related to unforeseen events such as non-adherence or emergencies. If participants failed to adhere to study guidelines or chose to withdraw, there had to be mechanisms in place to swiftly detect such instances. For high-risk patients, the REC advised maintaining constant contact to ensure early detection of any distress. Clear reaction protocols for unexpected events, like the identification of a risk factor or device failure, were needed. Typically, this would involve notifying the GP or a specialist, and relevant templates, such as a GP letter, and specified method of contact were requested. Participants should be informed of potential incidents and the related actions.

RECs were asked about staff and clinician training if it was missing from applications. They stressed the need for proper training, whether through written instructions, sessions with company reps, or on-site experts. Where applicable, proper sterilisation or disinfection of study materials and clear listing of device contraindications to avert potential risks were emphasized.

**Table 9.** Example quotations from meeting minutes on the ‘Harm Prevention’ subtheme, organised by codes.

| Code | Example Quote |
| --- | --- |
| Staff and clinician training | “Members requested confirmation that staff had been fully trained in the use of the device(s).” |
| Unforeseen events | “Please add more information into the [info sheet] about how... the research team would monitor the ECG alerts and recordings and contact them in the event of worrisome results.” |
| Non-adherence or withdraw | “The Committee questioned if the sensor showed a patient wasn’t complying, would the participants be switched to face-to-face therapy. The applicant clarified if they saw on the sensor that a patient hadn’t engaged with exercises within two days, this would trigger a call from the physiotherapist to identify the reasons[...] The outcome of the call would be recorded and if the patient hadn’t engaged after a further three days another phone-call would be triggered. If the physiotherapist felt the participant was struggling, then they would prompt a face-to-face meeting. The Chief Investigator also confirmed the participant would stay in the study. The Committee accepted this response.” |
| GP or specialist contact | “The Committee questioned how likely it would be the GP would respond. Moreover, under what circumstances would a GP be contacted and how would this be consistently applied; please provide clarification.” |
| Participant and co-habitant contraindications<br>Safety concerns | “Members explained to the applicants that it was important to inform participants that the wearable device (garment) should not be worn if there was an oxygen tank in the same room as this could cause a fire. Members...stated that anyone using oxygen in the house should be classed as an exclusion criterion, not just the participant.” |

### Theme 3: Digital Health Research Peculiarities

**Table 10.** Summary of discussion topics and outcomes under the theme ‘Digital Health Research Peculiarities’.

| Subtheme | n of action points | n of satisfactory responses | n of applications | Codes |
| --- | --- | --- | --- | --- |
| Necessity and Value of Research Goals | 3 | 4 | 8 | Relevance of research aims to primary outcomes<br>Value and impact of interventions<br>Unique considerations in digital health<br>Shift in research focus<br>Direct beneficiaries and translation of research |
| Remote Encounters | 0 | 3 | 3 | Accuracy and consistency<br>Privacy concerns<br>Distraction and supervision<br>Adaptation post-COVID<br>Training and expertise<br>Communication clarity<br>Technical challenges |
| Technical Issues and User Challenges | 10 | 3 | 8 | Information disclosure<br>Participant understanding<br>Technical term clarification<br>Device mechanics and app functionality<br>Cautionary procedures<br>Technology details<br>Privacy concerns |

#### Necessity and Value of Research Goals

RECs sought clarity on the necessity and value of research goals. These topics emerged because of nuances inherent to digital health interventions.

A common theme among the RECs’ inquiries was the alignment between the stated research objectives and the proposed methods of assessment. Some study aims raised questions with unexpected chosen endpoints and a misalignment with traditional healthcare outcome measures.

The potential value and broader impact of digital health interventions were also a point of focus. The committees also sought clarity on the practical implications of the research to ensure that research findings were meaningful and translatable.

**Table 11.**
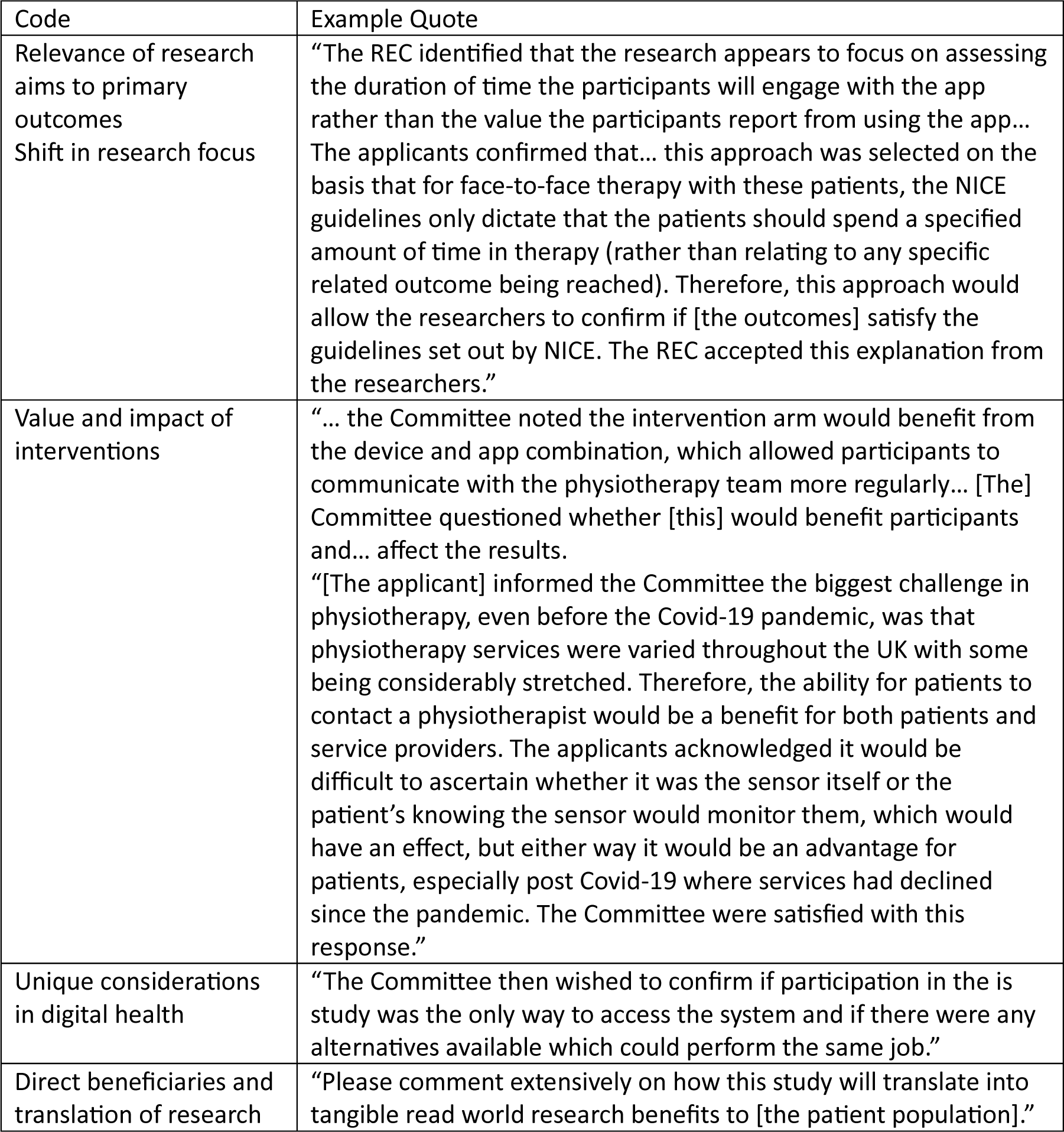
Example quotations from meeting minutes on the ‘Necessity and Value of Research Goals’ subtheme, organised by codes.

| Code | Example Quote |
| --- | --- |
| Relevance of research aims to primary outcomes<br>Shift in research focus | "The REC identified that the research appears to focus on assessing the duration of time the participants will engage with the app rather than the value the participants report from using the app... The applicants confirmed that... this approach was selected on the basis that for face-to-face therapy with these patients, the NICE guidelines only dictate that the patients should spend a specified amount of time in therapy (rather than relating to any specific related outcome being reached). Therefore, this approach would allow the researchers to confirm if [the outcomes] satisfy the guidelines set out by NICE. The REC accepted this explanation from the researchers." |
| Value and impact of interventions | "... the Committee noted the intervention arm would benefit from the device and app combination, which allowed participants to communicate with the physiotherapy team more regularly... [The] Committee questioned whether [this] would benefit participants and... affect the results.<br>"[The applicant] informed the Committee the biggest challenge in physiotherapy, even before the Covid-19 pandemic, was that physiotherapy services were varied throughout the UK with some being considerably stretched. Therefore, the ability for patients to contact a physiotherapist would be a benefit for both patients and service providers. The applicants acknowledged it would be difficult to ascertain whether it was the sensor itself or the patient's knowing the sensor would monitor them, which would have an effect, but either way it would be an advantage for patients, especially post Covid-19 where services had declined since the pandemic. The Committee were satisfied with this response." |
| Unique considerations in digital health | "The Committee then wished to confirm if participation in the is study was the only way to access the system and if there were any alternatives available which could perform the same job." |
| Direct beneficiaries and translation of research | "Please comment extensively on how this study will translate into tangible real world research benefits to [the patient population]." |

#### Remote Encounters

There was a low frequency of concerns surrounding remote encounters, and none led to action points. However, these issues merit attention for the unique challenges posed by the shift to digital formats, particularly following COVID-19.

The accuracy and consistency of remote assessments had to be confirmed, especially when comparing in-home versus hospital settings. When sensitive questions were posed, participants’ privacy during remote interviews had to be protected. Another point of interest was the potential distraction and supervision challenges during online interviews, particularly when young children were involved.

**Table 12.**
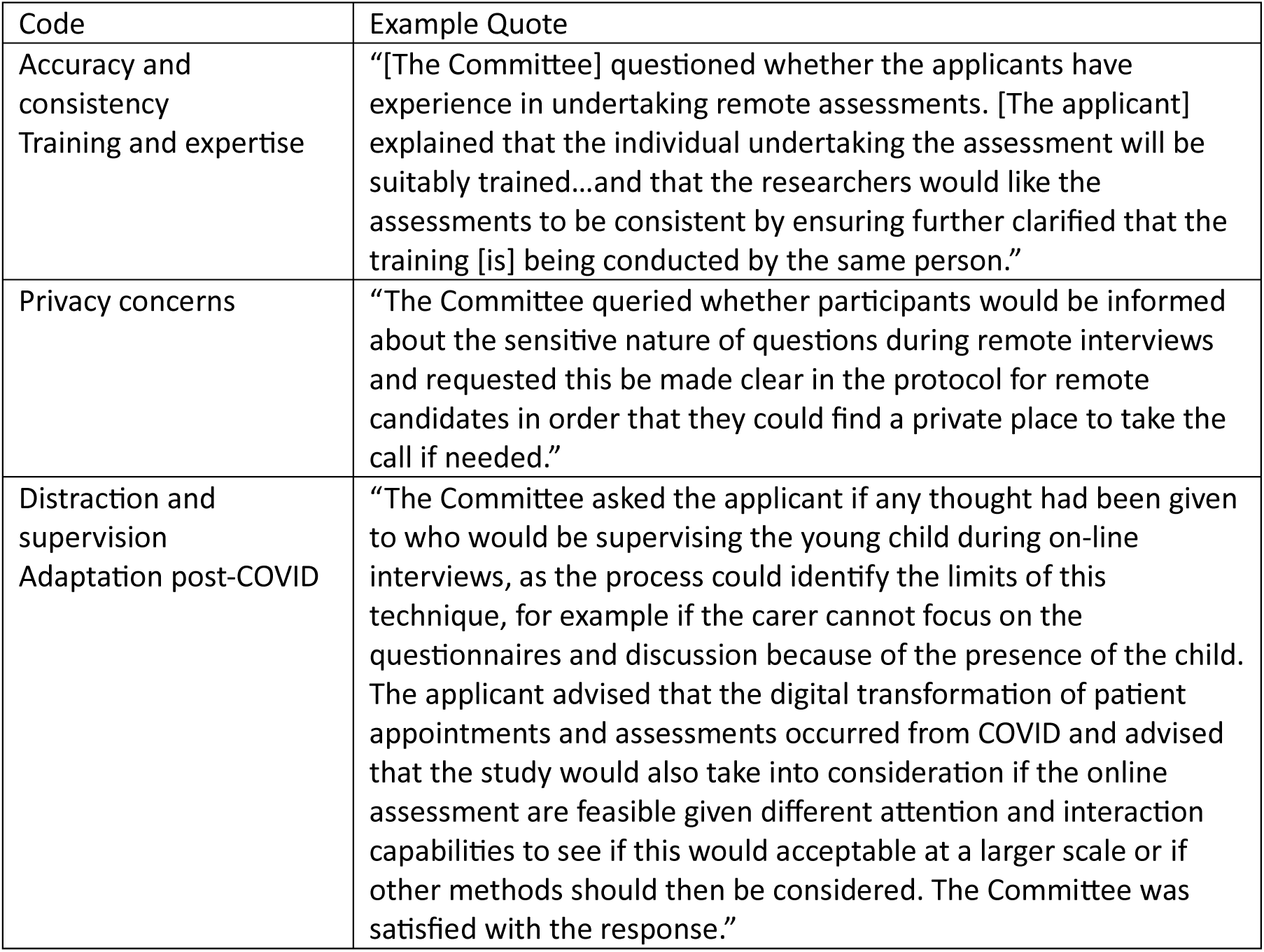
Example quotations from meeting minutes on the ‘Remote Encounters’ subtheme, organised by codes.

| Code | Example Quote |
| --- | --- |
| Accuracy and consistency<br>Training and expertise | "[The Committee] questioned whether the applicants have experience in undertaking remote assessments. [The applicant] explained that the individual undertaking the assessment will be suitably trained...and that the researchers would like the assessments to be consistent by ensuring further clarified that the training [is] being conducted by the same person." |
| Privacy concerns | "The Committee queried whether participants would be informed about the sensitive nature of questions during remote interviews and requested this be made clear in the protocol for remote candidates in order that they could find a private place to take the call if needed." |
| Distraction and supervision<br>Adaptation post-COVID | "The Committee asked the applicant if any thought had been given to who would be supervising the young child during on-line interviews, as the process could identify the limits of this technique, for example if the carer cannot focus on the questionnaires and discussion because of the presence of the child. The applicant advised that the digital transformation of patient appointments and assessments occurred from COVID and advised that the study would also take into consideration if the online assessment are feasible given different attention and interaction capabilities to see if this would acceptable at a larger scale or if other methods should then be considered. The Committee was satisfied with the response." |

#### Technical Issues and User Challenges

Other than patient safety and support topics covered above, most action points in this category revolve around ensuring participants have a comprehensive understanding of the digital tools employed. Some questions around the mechanics and functionalities of a device or app were raised.

**Table 13.** Example quotations from meeting minutes on the ‘Technical Issues and User Challenges’ subtheme, organised by codes.

| Code | Example Quote |
| --- | --- |
| Technical term clarification | "The Committee noted that the word "robotic" had appeared several times in the documentation and wished for an explanation as to what was meant by the word 'robotic'." |
| Cautionary procedures | "Mention whether sleep difficulties will be likely when wearing the technology." |
| Technology details<br>Participant understanding | "In the Participant Information Sheet... the Committee advised the applicant added a comparison between the risks and the benefits to using the device in contrast to the other procedural alternatives." |
| Information disclosure<br>Device mechanics and app functionality<br>Privacy concerns | "The Committee queried whether the device would pick up additional sounds or telephone conversations in the background. [The applicant] explained that this would be constantly reviewed as privacy was potentially a problem. [He/she] stated that there were two microphones on the device and... that additional sound could not be cancelled out until the researchers received the device data. [He/she] stated that he could put some information in the Participant Information Sheet about this." |

### Theme 4: Data Governance

**Table 14.** Summary of discussion topics and outcomes under the theme ‘Data Governance’.

| Subtheme | n of action points | n of satisfactory responses | n of applications | Codes |
| --- | --- | --- | --- | --- |
| Data Storage and Access | 11 | 11 | 16 | Storage on personal devices<br>Physical storage<br>Retention duration<br>Data access<br>Participant withdrawal<br>Regulatory compliance<br>Non-team access |
| Data Transfer and Sharing | 8 | 5 | 9 | Third parties<br>International involvement<br>Data transfer<br>Cloud and network |
| Anonymisation and Confidentiality | 7 | 1 | 6 | Identification protocols<br>Confidentiality measures<br>Data retention clarity<br>Pseudonymisation vs. anonymisation |
| Video and Audio Recordings | 4 | 3 | 6 | Selective recording<br>Data deletion post-processing<br>Privacy concerns |
|  |  |  |  | Transcription procedures<br>GDPR & data minimisation<br>Purpose of recording |

#### Data Storage and Access

About 60% of applications faced queries around data management, with focus on data storage, access and transfer. When action points came up, most were not of dire impact. Amendments caused delays in the approval process, but were not frequently the primary drivers behind provisional opinions.

Concerning storage on personal or company devices, applicants were prompted to de-identify data, clarify device ownership, and justify the use of unconventional devices like memory sticks and their safeguard measures. For physical storage, RECs asked about alignment with NHS standards and security measures at data storage sites. If not proposed, RECs requested that data should not be kept indefinitely and any unneeded data should be deleted.

If not specified, RECs inquired about the handling of data when a participant withdraws or is unable to participate post-screening. Applicants also grappled with complexities such as removing data that had already been de-identified.

**Table 15.**
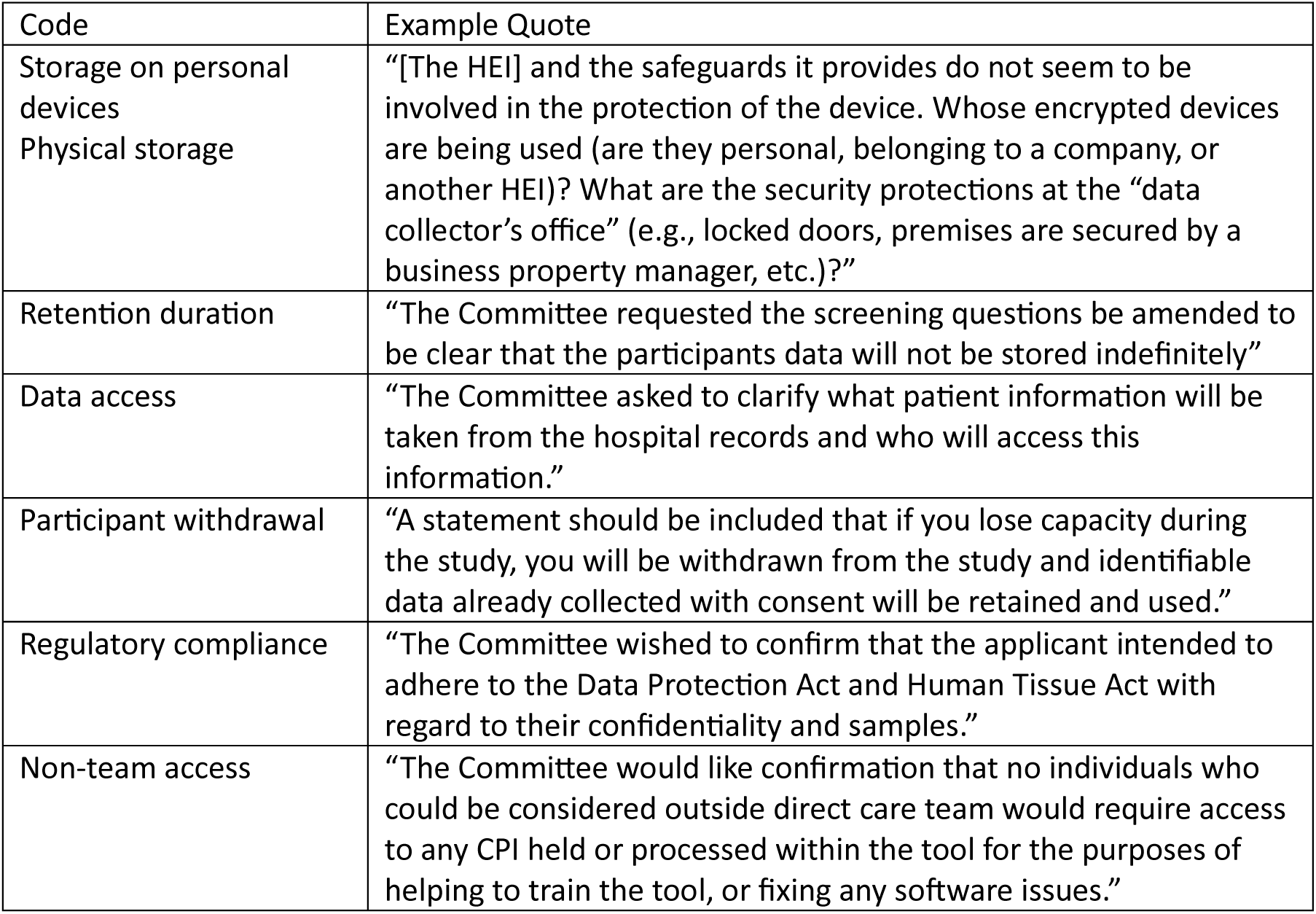
Example quotations from meeting minutes on the ‘Data Storage and Access’ subtheme, organised by codes.

#### Data Transfer and Sharing

RECs requested more information on who will be accessing data beyond the research team, such as during training and maintenance. Studies involving non-local collaborators often needed to clarify governance procedures and justify data storage on the cloud and transfers to the network or outside the UK. Details on how transfers between NHS servers, universities, and other collaborators would be kept safe were needed. Data transfer to commercial entities or third parties raised questions on access rights, GDPR compliance, international collaboration permissions, third-party data security, and the decision-making process for data storage locations, especially when HEI collaboration was involved.

**Table 16.** Example quotations from meeting minutes on the ‘Data Transfer and Sharing’ subtheme, organised by codes.

| Code | Example Quote |
| --- | --- |
| Data transfer | "The Committee queried how data would be transferred between NHS servers, [the applying enterprise] and the [HEI]." |
| Cloud and network<br>Third parties | "A further clarification was requested in relation to the data storage, security and tracking features and the location data collected by the Apple watch.<br>"[The applicant] explained that the location data is not collected as part of the study and it can be turned off completely should the participant wish to do so, as it is not required for the app to work. All the data is stored on [a cloud service] servers which are password protected only accessible by authorised staff. The Committee was content with the explanation. " |
| International involvement<br>Cloud and network | "Please can you clarify further how data is processed outside the EEA but not stored? Is it that the server of the cloud is based in [a large city in the UK]?" |
| International involvement<br>Data transfer<br>Third parties | "The Committee asked for clarification as to who would have access to data when it was transferred to the commercial company. The Chief Investigator assured the Committee they were fully GDPR compliant and no one would have access to the back-end data except for the Chief Investigator and one person in IT. The researchers elaborated that there was a dashboard which provided metric data which was shared with the commercial company. However no specific personal data was contained in this. Furthermore, the Chief Investigator assured the Committee they would not provide any data collected in the UK to Canada or US, and vice versa. The Committee were satisfied with this response." |

#### Anonymisation and Confidentiality

RECs sought explanations for the use of identifiable information, such as a patient’s name on forms, questioning if codes or numbers could be preferable. RECs highlighted the importance of distinguishing between pseudonymous and anonymous. Detailed strategies of anonymisation and how confidentiality would be maintained were requested when missing. UK Data Service and ICO were recommended to applicants.

**Table 17.** Example quotations from meeting minutes on the ‘Anonymisation and Confidentiality’ subtheme, organised by codes.

| Code | Example Quote |
| --- | --- |
| Identification protocols | "Please provide explanation / justification why the patient's name is required on the interview / questionnaire form. Alternatively, a number or code would be preferable." |
| Confidentiality measures | "Provide more information and what steps will be in place to prevent the following (as stated in... the IRAS form): 'Although care is taken to ensure confidentiality, there is a possibility that participant's name or other personal information could be seen by an unauthorised person'." |
| Data retention/usage clarity | "It is not sufficiently clear what the 'app usage data' is and is not before and after anonymization, and whether this would be linked to either app account identifiers (if they exist) or survey responses before or after anonymization." |
| Pseudonymisation vs. anonymisation | "It is not sufficiently clear what the 'app usage data' is and is not before and after anonymization, and whether this would be linked to either app account identifiers (if they exist) or survey responses before or after anonymization." |

#### Video and Audio Recordings

Committees questioned the need and duration for video storage, asking for justifications. Often, when the applicant had no intention of recording, they would neglect to mention this in the application. This is shown by the frequent response assuring that no recording will be kept when committees sought clarification on whether interactions, like teleconferences, were recorded.

The need to share recordings, especially with sponsors, was a concern. In one case, skepticism existed regarding companies deleting valuable research data. (The applicant, who was interviewed, elaborated on this point, feeling a sense of distrust and expressing frustration towards the REC, as discussed later.)

**Table 18.**
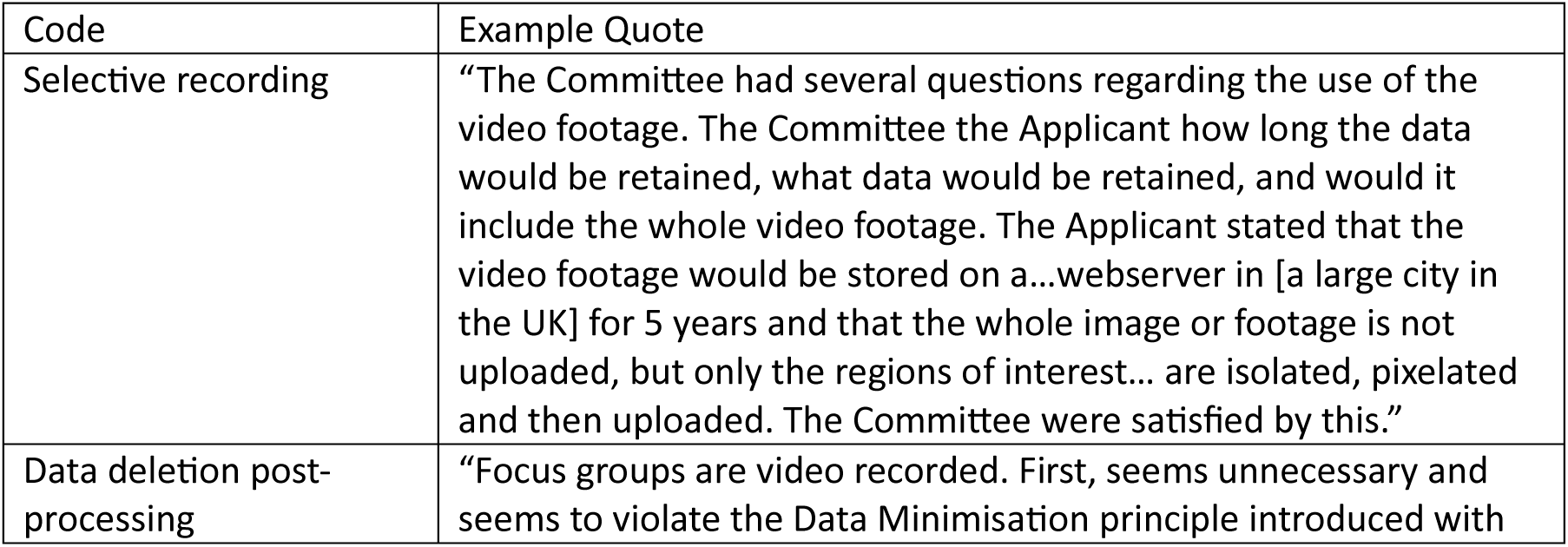
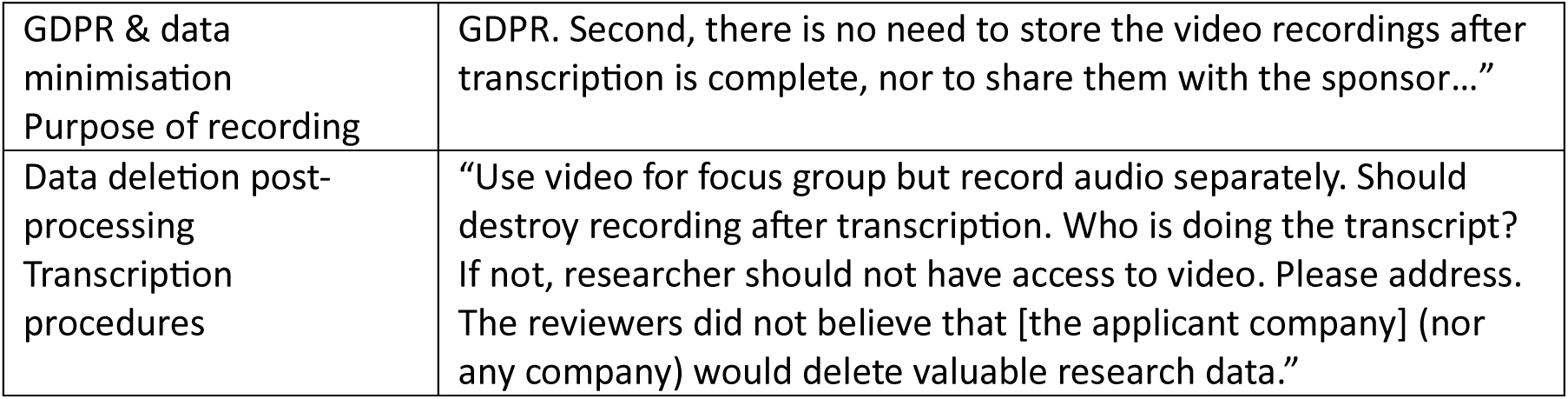
Example quotations from meeting minutes on the ‘Video and Audio Recordings’ subtheme, organised by codes.

### Theme 5: Document Quality and Completeness

**Table 19.** Summary of discussion topics and outcomes under the theme ‘Document Quality and Completeness’.

| Subtheme | number of action points | number of satisfactory responses | number of applications | Codes |
| --- | --- | --- | --- | --- |
| Insufficient Content and Detail | 87 | 4 | 25 | Clarifications or missing information<br>Background and context<br>Contact information<br>Data governance procedures<br>Study protocol details<br>Compensation, incentives, and cost<br>Conflict of interest disclosure<br>Consistency between information sheet and consent form<br>Ethics review information<br>Participant safety and support<br>Purpose of the study<br>Useful images |
| Missing Required Documents | 15 | 0 | 11 | Needing additional consent form and information sheet<br>Researcher credentials<br>Document standards<br>Training evidence<br>Prescriber contact documents<br>Advertisement and invitation clarity<br>Clinical trial registration<br>Device specific information<br>Miscellaneous documents |
| Accuracy and Presentation | 55 | 2 | 20 | Wording adjustments<br>Lay language use<br>Formatting enhancements<br>Template precision<br>Irreverent sections in template |
|  |  |  |  | Avoiding inaccuracies and redundancies<br>Reader-friendliness<br>Consistency across documents<br>Grammatical errors and typos |

#### Insufficient Content and Detail

The most common change needed was the addition or editing of information omitted from participant information sheets or consent forms, mostly covering topics from previous themes. These usually do not result in long discussions, but led to a high number of action points in over 80% of applications. Other than fixing simple omissions, many were a result of discussions covered under previous subthemes, requiring amendments on participant-facing documents. Some information included in the information sheet needed to be added to the consent form, and vice versa. Examples of common easy-to-fix omissions include ethics review information, contact details, and purpose of the study. Where applicable, applicants may be asked to include instructions with images or pictures of the device for visual guidance.

**Table 20.** Example quotations from meeting minutes on the ‘Insufficient Content and Detail’ subtheme, organised by codes.

| Code | Example Quote |
| --- | --- |
| Background and context | "A line should also be added to the beginning of the lay summary to briefly describe the disorder and should also state that the study includes children from age three and adults lacking capacity to consent." |
| Consistency between information sheet and consent form | "The Committee highlighted the information sheet states, 'We will let you know the results of the study when it is finished...' This should be reflected on the consent form." |
| Contact information | "Please consider adding contact details for complaints" |
| Data governance procedures | "Under the heading, 'What will happen if I do not want to carry on?' The Committee agreed this section should be updated to state participants have the right to withdraw data." |
| Study protocol details | "The Committee requested the PIS be updated to include text stating the reason they will ask about recreational drug use[...]" |
| Compensation, incentives, and cost | "The Committee requests clarification be provided regarding how the £50 compensation will be made, with this being clear in the PIS" |
| Conflict of interest disclosure | "The Committee highlighted the disclosure...regarding a potential conflict of interest... and agreed this should be outlined in the information sheet for full transparency to participants." |
| Ethics review information | "State that the London-Chelsea Research Ethics Committee has given a favourable opinion of the study." |
| Participant safety and support | "Add text to the speech language therapist PIS stating that deciding not to take part will not impact their job." |
| Purpose of the study | "Make clear that this is a pilot study." |
| Useful images | "Members of the Committee determined that the picture of the glasses in the protocol was very useful. Please update the information sheet to include this picture." |

#### Missing Required Documents

The RECs frequently noted omissions in the documentation provided. On multiple occasions, the committee requested CVs of research team members in a standardised format and evidence of training. Applicants had often missed out documents like invitation letters and emails, adverts, device instruction sheets, specific protocols, GP letters, and forms that would be used. Additional consent forms or sections are also needed for separate participant groups.

**Table 21.** Example quotations from meeting minutes on the ‘Missing Required Documents’ subtheme, organised by codes.

| Code | Example Quote |
| --- | --- |
| Needing additional consent form and information sheet | "A specific Participant Information Sheets/Informed Consent Forms should also be created to reconsent sixteen-year-olds who have capacity to consent or to obtain a declaration from their consultee." |
| Researcher credentials<br>Document standards | "The Committee commented that [a study team member] had a very long, detailed CV. The Committee wished for a version in line with the HRA's CV template. The applicant confirmed the two-page version would be sent." |
| Training evidence | "Please provide evidence of GCP training or equivalent" |
| Prescriber contact documents | "The Committee requested the GP letter be submitted." |
| Advertisement and invitation clarity | "The online advert provided states that it is example text. The Committee requests the final version of the advert be provided." |
| Device specific information | "The Committee agreed that a separate information sheet for each specific device would need to be created... Please provide the information sheets for [devices that have been chosen] and submit any future ones as substantial amendments." |
| Miscellaneous documents | "The Committee noted that a copy of the actual insurance certificate had not been submitted." |

#### Accuracy and Presentation

Many changes were requested on wording and formatting. Changes in wording were usually for the purpose of conveying messages more accurately or increasing reader-friendliness with lay language. Formatting changes were requested to improve document clarity and readability. RECs insisted that authority-provided templates be used precisely as given. However, when using study team-created templates, some sections were found irrelevant, leading to action points for editing. For example, a checkbox for “Yes” or “No” was requested next to all optional items on the consent form.

Changes requested during the meetings or oversight occasionally led to inconsistencies across different documents or versions not being up-to-date. Applicants were then required to remove irrelevant content to prevent inaccuracies and redundancies. RECs also pointed out grammar mistakes and typos.

**Table 22.** Example quotations from meeting minutes on the ‘Accuracy and Presentation’ subtheme, organised by codes.

| Code | Example Quote |
| --- | --- |
| Wording adjustments | "The Participant Information Sheet should be changed from 'Why have I been chosen? To 'Why have I been invited?'" |
| Lay language use | "Please check the text in all participant-facing documentation for technical or non-lay language and either remove or reword." |
| Formatting enhancements | "Please consider making the PIS easier to navigate by changing the headings of each new section to be emboldened to differentiate from the core text." |
| Template precision | "The HRA's GDPR wording has not been used verbatim – please can you follow the instructions of use on the website and update the PIS." |
| Irrelevant sections in template | "Please ensure it is relevant to the proposed project only and does not duplicate clauses from the challenge study." |
| Avoiding inaccuracies and redundancies | "The Protocol para 11.2 should be amended to remove reference to an interview and focus group." |
| Reader-friendliness | "The Committee agreed that the master PIS is quite long and dense. It includes some complex phrases and terms, and should therefore be revised to make it lay reader friendly." |
| Consistency across documents | "The Subcommittee noted that the consent form states data will be stored for 5 years but IRAS form states 10...they asked for clarification of which is correct and to amend the incorrect document." |
| Grammatical errors and typos | "The Committee agreed that the Information Sheet for parents includes several grammatical errors and spelling mistakes and needs a complete rewrite." |

#### Impact and Severity

The issues in the subthemes Conflict of Interest, Anonymisation and Confidentiality, and Video and Audio Recordings were some of the most impactful. Although these subthemes cropped up in a limited number of studies, the vast majority of studies that encountered these barriers were provided with a provisional opinion.

The least impactful subthemes were Sampling and Recruitment, Accuracy and Presentation, Insufficient Content and Detail, Protocol Robustness, and Technical Issues and User Challenges. Out of all the studies where these subthemes identified as issues, only about half received a provisional opinion. However, the vast majority of action points that came up fell into the former three subthemes. These issues consistently emerged but carried minimal weight against a favourable opinion. This is not to say that these barriers are not substantial, since action points delay the approval process. For example, requests to provide instructions on the technology are always required to be submitted as amendments.

#### Comparison between RECs

Perspectives and opinions can vary across different RECs. On one occasion, applicants received a request for an additional consent form and information sheet for young children from an HRA REC that provided an unfavourable opinion. They later sought a second review from a different HRA REC, who agreed that “these documents [could not] be used as it would be difficult to get a meaningful response” from such a young patient population and gave a favourable opinion with additional conditions.

HRA RECs held meetings with the applicants, which allowed them to address some issues on the spot, whereas the HEI RECs replied to applications in writing, requiring the applicants to also respond in writing. The HRA REC meetings were documented in neutral language with an observatory tone, whereas the HEI REC documents were more question-oriented and less neutral in tone. However, the difference might stem from the HRA REC meeting minutes being a form of documentation and the HEI REC’s response being a medium that asks similar questions as would be posed in meetings (Table 23). Both RECs sought attribution, with the HRA RECs desiring proper acknowledgment for their ethical review and the HEI REC seeking visible affiliation through the inclusion of the university banner.

**Table 23.** Example quotations from meeting minutes of HRA and HEI RECs compared.

| HRA REC Meeting Minute | HEI REC Response |
| --- | --- |
| “The Committee asked how consenting would be managed in light of any Covid restrictions.” | “Why have signed consent forms – why not handle through REDCAP?” |
| “The Committee wished to confirm if there had been training for study doctors on how to use the device and if there would be a company representative on site.” | “Please comment extensively on your relevant experience for conducting this extremely sensitive study.” |

### Interview Findings

#### Enterprise and Study Characteristics

Table 24 summarises the characteristics of projects discussed during the interviews. Some interviewees talked about multiple ethics applications for different digital health interventions. One interviewee had provided assistance with the application process but was unaware of the approval status at the time of the interview, having ceased involvement earlier. One interviewee from a UK-based office of a global enterprise shared their experience with a US institutional review board (IRB).

**Table 24.** Summary of key attributes of interviewed enterprises and their projects.

|  | Micro to Small<br>Size Enterprise<br>(Up to 50 employees)<br>n=5 | Medium Size<br>Enterprise<br>(Up to 250 employees)<br>n=3 | Large Enterprise<br>(Over 250 employees)<br>n=2 |
| --- | --- | --- | --- |
| Intervention Type |  |  |  |
| Remote care delivery | 2 | 1 | - |
| Remote monitoring and management system | 3 | 2 | 1 |
| App for self-monitoring and information tracking | 0 | 1 | 1 |
| Approval Status |  |  |  |
| Approved | 2 | 3 | 2 |
| Pending | 2 | 0 | 0 |
| Unknown | 1 | 0 | 0 |
| Intervention Availability |  |  |  |
| Direct to consumer | 1 | 1 | 1 |
| NHS commissioned, available through practitioner | 4 | 0 | 1 |
| Available to employees through employer | 0 | 2 | 0 |

#### Common Barriers and Applicant Attitude

Interviewees recognised the important role of ethical approval in safeguarding participants during the study and as further developments arise from the research. When asked about the biggest barriers encountered in the application process, we identified six themes from the interviewees’ responses, detailed below.

#### Submission and Protocol Revisions

Interviewees’ frequent reports of edits in wording, grammar, and clarity mirrored the document analysis. Though interviewees were surprised by the emphasis on minute details, many felt such changes had limited repercussions.

**Table 25.** Example quotations from interviews with enterprise representatives on ‘Submission and Protocol Revisions’.

| Code | Quote |
| --- | --- |
| Submission clarity | "For this study, there were exploratory outcomes [measured through blood samples]. [The REC] had all these questions about how [it was] being labelled, is it being destroyed... [They] seem to want a lot of detail behind that, which I just didn't feel was particularly relevant." |
| Accuracy and presentation | "I think I was surprised to see that... grammar was important. Like I know that's necessary, but like stuff like the font used... I thought it'd be a bit more clinical." |
|  | "[The REC] said the [Terms and Conditions] are too long. We needed a summary. So all of this..., you then have to change specifically for a piece of research." |

Interviewees modified template wording, like consent forms and GDPR text. However, RECs favoured strict adherence to templates. Contrarily, one submission faced revisions for resembling a prior approved application. Some believed the lengthy REC-styled documents and perceived bureaucracy deterred patient involvement and site recruitment.

**Table 26.** Example quotations from interviews with enterprise representatives on ‘Submission and Protocol Revisions’.

| Code | Quote |
| --- | --- |
| Wording and templated material | "We cannot predict how many clinical appointments [patients] will have. They could have two or three during the study duration... We put there will be a few study visits[...] We couldn't guarantee to patients [a specific number of appointments], ...but [the REC] wanted something more specific, so we have to work with them to figure out the correct wording for it." |
|  | "[Something] I found very unhelpful was [that] the participant information sheets are very rigid and boring. We created an onboarding pack. The service designers [spent] a long time working with people... from a patient group to make sure things that were designed and language... would resonate with them. [The REC stated] that it has to be within the confines of the participant information sheet template and it has to be... this font. And you can't use any colours. ...Very frustrating. So, we...[packed] all of this information into...an 18-page... information sheet that was totally unreadable.<br>"It couldn't deviate from a long... Word document. It had to be dry and... factual language.<br>"We had a lot of complaints from GP practises who are trying to implement the service. They had to give this to patients and patients that found it very, very difficult to absorb all of this information." |
|  | “We ended up signing 26 of these contracts, ...which was challenging. You have to find the person who can legally sign, [get] all their information, then send them [this long] document with confusing legal content. From an admin perspective, it was a nightmare.” |
|  | “The consent form [we submitted] wasn’t explicitly the template that they put online. The points... weren’t the exact same, but obviously I didn’t want the plagiarise their consent form, so I changed some of the words, but we weren’t allowed to do that.<br>“When you’re writing your own... information sheet for your own study, they are meant to be individualised per study.<br>“[In] the exact one, some of the points weren’t even relevant to my [study], but they said we want you to put it in anyway. I was like, ‘right, OK, I’ll do it.’ So, we did that and that was probably the hardest point. That was back and forth for a couple of months.<br>“I don’t know because... my [team member] used to work for IRAS, and [he/she]... couldn’t believe how fussy they were being. It wasn’t even in terms of the actual study..., it was the formatting.” |

Study design revisions presented challenges, with consent protocols and data governance perplexing to justify and mitigate. Interviewees sought clearer guidance. Table 27, example 6 revisits the scenario from Table 18, example 4, offering the enterprise’s perspective. The interviewee’s stance is elaborated in the following section.

**Table 27.** Example quotations from interviews with enterprise representatives on ‘Submission and Protocol Revisions’.

| Code | Quote |
| --- | --- |
| Consent | “The traditional consent process[...][requires] a patient information leaflet [and] 48 hours to think about it. Given our intervention—a phone call for 8 minutes—it was challenging[...] It was not feasible to collect physical consent because of [multiple] surgical locations[...] So we [had] to make an ethical trade-off: either ‘gold standard’ consent... and exclude many patients, ...or phone-based consent. After much deliberation[...] we decided [on] phone-based consent, ...which was well received by the Ethics Committee.<br>“The ethics process... has a clear default way to consent patients and it’s set out by non-digital health studies[...] If you’re concerning to a gene therapy that’s being tried for the first time, you absolutely need to read the fine print and wait two days before making up your mind. But [for] an additional phone call, ...it’s probably okay to have that discussion on the phone. For us, that was ...frustrating having to justify that.” |
|  | “Guidance around...commensurate consent would have been really helpful... Different things will have different ethical challenges, so I don’t think there’s a way to get away from the individual nature of the evaluation, but I think...the guiding principles would have been would have been helpful.<br>“[The HRA] had a useful document around proportionate consent. This was not quite applicable to us; it was an e-consent statement.” |
|  | “How patients are informed...and consented is very antiquated and not suited for digital health products. The default is still a piece of paper with four or five pages [of something like] terms and conditions.<br>“By having this crazy consent process, you actually skew...and create performance bias in the study.” |
| Data governance | “For some very strange reason, instead of seeing the therapists as... [members of] the research team, ...they were [viewed as] participants. Our therapists had to sign a consent form. And [while] they were going to contribute to the authorship, training, and... [evaluation] of the AI, [the committee] insisted that they were participants and that they had to be kept anonymous. We knew their names because they were part of the author team.<br>“It was the strangest thing ever. So completely baffling, but that was a requirement from ethics... so I have to comply, doing very strange things to keep their names anonymous. It delayed the research by, I would say, two months.” |
|  | “They were very concerned about [us] being able to access the data that we got out of patients. I didn’t fully understand why, because it’s our software platform. We can’t not have access to the data because sometimes you have to do something with the data. This idea [of making] it truly anonymous, it just doesn’t work with the way software platforms are configured.” |
|  | “It was so difficult because of...the ownership of the data. Protecting the patients’ data rights [in relation to] a piece of research which is trying to help them was a challenge. We couldn’t get through ethics because they were [concerned about how] the data was being held, what data we were holding, and [whether] we really needed to collect [and keep] it. They were so protective of the patient that they made it almost impossible for us to deliver a solution[...] We work with [a large number of] NHS trusts in the real world. Why [is it that] in this research environment, [they imposed] so many constraints?<br>“In digital health, a company [provides] a service and [does] some research on it. The data...tends to be hosted within the organisation providing the service. That makes sense. Sometimes we have projects denied initially because [RECs] wanted us to have the data for our own users, our own, service users hosted in the [HEI].” |

#### Dynamic between Parties

Relationships between research teams, sponsors, REC, HEI, and external partners, while often supportive and collaborative, can be complex or hindering. Interviewees expressed hesitancy to engage with RECs and feeling a sense of distrust from the RECs. Table 28, example 1 shows the interviewee’s frustration following example 6, Table 27.

**Table 28.**
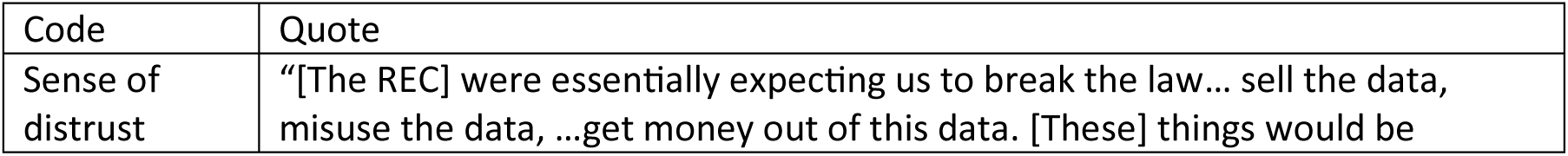
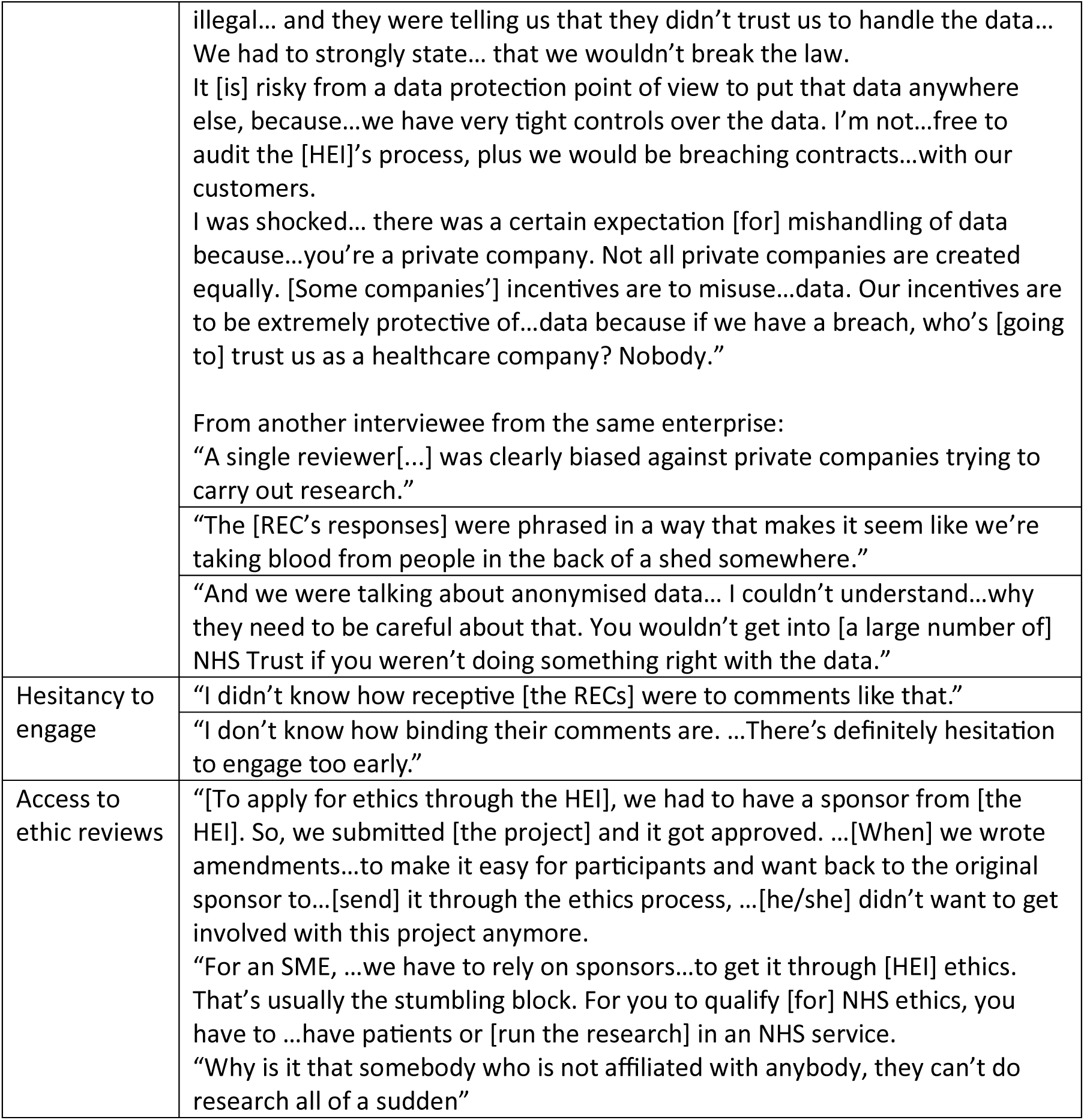
Example quotations from interviews with enterprise representatives on ‘Dynamic between Parties’.

#### Application Time and Procedures

Interviewees reported the ethics application and the steps leading up to it as time-consuming and effort-intensive. Many had not anticipated the length of the process, and those who did often sought to avoid ethics approval if possible.

**Table 29.** Example quotations from interviews with enterprise representatives on ‘Application Time’.

| Code | Quote |
| --- | --- |
| Steps leading up to the application | "You have to... go through a two-month process of putting together a protocol. Then...wait for two to three months while somebody [peer] reviews it... Then through IRAS. And those things...can't be done at the same time. |
|  | "I was very surprised to learn how long it would take... By the time the project finished, [collaborating partners] had completely lost interest...and were moving on to another therapeutic area." |
|  | "Clarification [with the REC] around whether or not we actually [needed] to get research ethics approval...took quite a while. It was mostly all done over phone." |
|  | "You've always got a queue and...they never go fast enough." |
| Long process and response time | "Usually, ethics applications take...months or so. We try to avoid having to do ethics applications if we can because we know that can...significantly delay the process." |
|  | "I think it really comes down to the fact that the digital health industry moves very quickly, and generally, IRBs move slowly," |

Interviewees encountered difficulties with the IRAS portal, and factors typically insignificant in traditional research became obstacles in digital health research. The relevancy of application questions was an international issue, and submitting even minor changes proved problematic and stressful.

**Table 30.** Example quotations from interviews with enterprise representatives on ‘Application Procedures’.

| Code | Quote |
| --- | --- |
| IRAS usability | "It's not intuitive to use... It's not particularly user friendly." |
|  | "I don't think that the guidance they provided was extensive enough... I phoned up the IRAS team and they didn't know how to fill in the form." |
|  | "I don't get [a status update] that it's being reviewed. Only when it was approved." |
|  | "Everything else is left centred and [question] A68 is a drop-down box... It's up on the right on its own. [If you don't fill in A68], [the system] won't let you submit it, so it puts a delay in preparing the form... [The system] says 'You haven't completed the form' and you're looking and looking for the question you haven't answered... I've done it 10 times and I still miss it." |
| Content relevancy | "[The system] is clearly designed for drug trials. [There are] some minor tweaks for clinical investigations of medical devices[...] You put in N/A in load of boxes that...aren't relevant." |
|  | "I think the institution primarily serves clinical trials, drug trials... for academia communities. And because the digital health industry is still up and coming, there's...sections that aren't...applicable." |
|  | "I don't believe there are particular areas in the application that apply to only digital health... I think that a lot of IRBs can be a bit more up to date" |
| Change submission | "[When] I finished the [requested] amendment, the biggest barrier was [getting] the signatures on the IRAS form... The IRAS site sends a personalised e-mail out to every collaborator on the study, which is 7 people [for my study]. ...I accidentally forgot to rename a version number... And if you accidentally touched a button, they all [get] wiped. You have to resubmit the amendment within 24 hours and getting everyone's signatures [again]. I've had to do the signature thing four or five times...It was just really buggy." |
|  | “We had to make a very minor update... But updating and going back through is a process that fills people with dread... And my feeling...was that it was not very clear how long that takes. In the end, I think it was very straight forward actually. It turned around in a month. ...I remember the study team being really stressed about that.” |
| Technological flexibility | “It’s very difficult to make changes and get it right first time. For digital health and for technology in general, especially software, you want to iterate and you want to be a bit more flexible.<br>“We thought we had to freeze the algorithm. We didn’t make any major software updates that were not safety critical. I would like to do a study soon using generative AI. Because it’s a new technology, ...we would want the ability to tweak the system...and update the algorithm to [patient] response. And I’m not sure how that will be received by the HRA.” |
|  | “The app is constantly iterating. Features are changing[...] These small amendments... are costly.” |

#### Acumen and Practicality in Digital Health

Given the novelty of digital health research, RECs sometimes miss its nuances, leading interviewees to clarify technicalities and face unrealistic amendment requests. While acknowledging the importance of inclusivity, interviewees were frustrated by its practical challenges. Similar issues arose regarding data governance in the Submission and Protocol Revisions subtheme.

**Table 31.** Example quotations from interviews with enterprise representatives on ‘Acumen and Practicality in Digital Health’.

| Code | Quote |
| --- | --- |
| Familiarity with Digital Health Technology | “The biggest challenge was...trying to explain what it did in the session... I doubt it was a familiar concept to many people in the NHS at the time.” |
|  | “People tend to have less knowledge about digital health, ...but I’m sure that people in other disciplines feel similarly.<br>“For example, the apps...store information in a private server in the back end. That doesn’t go to [the HEI] ... [or] my laptop. [The REC was] getting confused and thinking it was going to my laptop... [or] an e-mail address..., so I had to clarify quite a lot of that.” |
| Practicality and monetary constraints | “[The committee] asked us... to include more different types of study phone... We currently have up to the iPhone 14, but if another new phone comes out, they wanted a note on it saying that any phones can be calibrated [to use] the... app.<br>“There is no guarantee that we’ll be able to do it within the time, because calibrating the app to each phone takes a crazy amount of time. But they said... you can’t exclude a participant because they don’t have a certain type of phone.” |
|  | “This issue of inequalities is a massive one. But you are really doing something for 0.5% of people where that is an issue, versus the 99.5% [when] you can’t even deliver something basic to them... I know that I’m not supposed to think |
|  | like that, but there are times you do think like that... When the system doesn't have the money to do [the basics], you want to throw more challenges in?<br>"There's a general need to determine when ethics... need to be more pragmatic [or] absolute, ...recognising that it's more important that the research is done... If I'm a patient with...cancer, if I can get anything that's [going to] help [me], I'll give my data." |

#### Support and Guidance from RECs

Interviewees contacted the REC and HRA for support and consulted online resources published by the HRA during the application process. Multiple interviewees reported that the REC/HRA was unsure of the answer to their questions. The timeliness of responses varied across different RECs and experiences.

**Table 32.** Example quotations from interviews with enterprise representatives on ‘Support and Guidance from RECs’.

| Code | Quote |
| --- | --- |
| Support<br>timeliness | "You don't get assigned to a REC until after you submit everything. We had a lot of questions before we submitted things, which we asked the HRA... There [were]...a lot of the questions they couldn't answer. It took a while to get to the right person, but their [point of contact] [was] reasonably responsive." |
|  | "The biggest challenge was... lack of timely support. There's no real contact... Sometimes it's really useful just having someone to phone, but you're just left with the non-replied e-mail of doom." |
|  | "[REC X] is very good. [They] got back to me [in] two days. Then [REC Y], one to two weeks. [REC Z] [...]I didn't get a response for three months there. "[REC Z] has a study wide form... I can't do any work in the [RECs X and Y] until that's released. [REC Z] is blocking all three trusts." |
| Expectations<br>and<br>transparency | "It was not very clear how long [the amendment] takes." |
|  | "I don't even know how people actually checking the answers... [on the application form] because you don't really get any feedback." |
| Support<br>quality | "[The REC] [didn't] know anything about the IRAS form. For example, ...my study was on a phone app. I didn't know whether to [submit] it as a clinical study, a clinical validation study, or a medical device study, because it is a CE marked device, but it's not been used in a clinical trial... If I put it in as a medical device, it's going to charge me £4000 to get MHRA referral?" |
|  | "We [asked] whether or not we actually need to get REC approval... They basically said we don't really know. So just do it." |

#### Enterprise Expertise and Resources

Expertise in research ethics varied among enterprises, from novices to seasoned experts. Some had internal regulatory expertise while others relied on academic collaborators. While many faced similar challenges, enterprises with extensive ethics experience often reported a more positive or neutral application experience.

There’s a recognised need for targeted resources; while HRA resources were appreciated, gaps were identified. Interviewees also consulted various healthcare and technology guidelines, frameworks from EMEA and the FDA, online resources published by HEIs, and internal enterprise materials.

**Table 33.** Example quotations from interviews with enterprise representatives on ‘Enterprise Expertise and Resources’.

| Code | Quote |
| --- | --- |
| Resources consulted | "We didn't particularly have [expertise internal to the company]. It's mostly [HEI] people [we consulted]." |
|  | "I've seen...webinars on procurement and selling. But I don't think I've seen...an event [on] 'how can industry do research'. ...I'm sure there is [information] if I [really searched] for it, but none that's readily available." |
|  | "[I referred to] the Digital Technology Assessment Criteria (DTAC) for user experience design. ...I know a lot [about] how digital health technologies...get approved for use within the NHS and for the government, so I applied all those things, made sure that there's a lot of patient engagement, user feedback." |
| Varied experience and knowledge | "I didn't know that a company could independently submit an ethics application. I thought it had to be researchers [from academia]." |
|  | "Some people are better at getting through ethics than others. That has a huge variability in it. I've noticed that some academics really understand this. Some don't... That's the advantage of working with the HEI, ...whereas we'd be clueless...if we [tried] to do it ourselves." |
|  | In response to the interviewer's question on protocol development:<br>"If you're at a startup, often lots of this stuff just isn't done because people don't know about it, which means they learn quickly, but can often have lots of holes." |

#### Common Enablers

Support, particularly through feedback from the REC and expertise accessed through various channels, was a primary enabler. PPI guidance bolstered protocol justifications and material development. One interviewee mentioned the benefits of connecting with local colleagues informally and advised an open mindset regarding reservations around sharing information: “Don’t be fearful. Just be careful.” Many interviewees emphasised the importance of a detailed protocol for team alignment and submission preparation, with templates and past examples being particularly valuable.

#### Suggested Improvements

Interviewees voiced desired changes regarding the challenges they faced during the application process, detailed below.

**Table 34.**
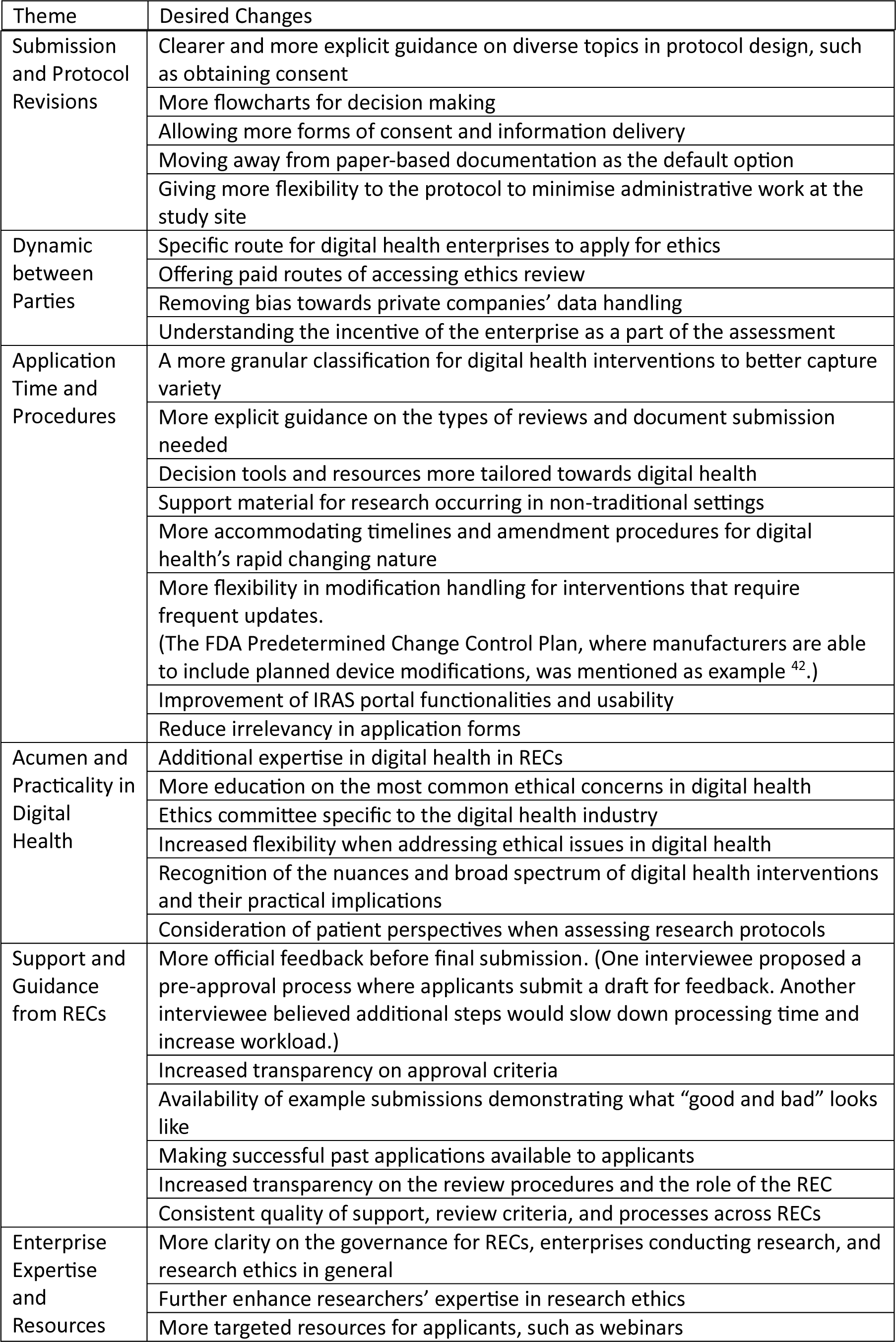
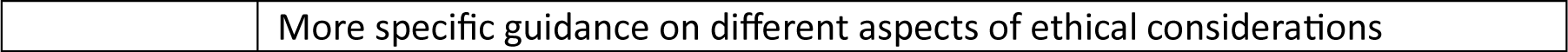
Desired Changes Voiced by Interviewees Based on Challenges Faced, Organised by Associated Themes.

#### International Experience

An interviewee shared their experience with a commercial North American institutional review board (IRB), chosen for specific reasons by the enterprise. This IRB offers ethics reviews for institutions without their own boards and provides consulting and training services. The interviewee identified enablers including a user-friendly application portal, responsive customer support, and quick turnaround times. However, the primary challenge was the high application costs, including fees for initial submissions and amendments.

## Discussion

We found a complex picture of the challenges in the digital health ethical landscape. The ethical review process demands considerable time and effort, irrespective of experience. For newcomers, it can be particularly daunting, with a steep learning curve.

The findings emphasise the importance of providing detailed and high-quality application documents with well-developed study designs. Comprehensive protocols with arrangements for potential concerns are key to mitigating substantial amendment requests. Study designs and objectives should be justified with clear reasoning rooted in evidence, established guidelines, or PPI recommendations. A unified protocol embraced by the entire team is crucial, as is their preparedness for meetings where this protocol is presented. A participant-centric approach should be taken when preparing the application, making sure documents are accurate, up-to-date, avoid jargon, offer tailored support based on the intervention’s complexity and sensitivity, and allow participants adequate time and space for consent and engagement.

While some barriers require greater attention, proactive effort can minimise post-submission delays. In research involving vulnerable patient groups, topics like inclusion criteria and protocol rigour invariably require in-depth discussions, often necessitating reference to regulations or care standards. Addressing such inherently multifaceted issues demands additional funding, extended discussions, and thoughtful deliberation. Substantial modifications to the study’s design may be required in the absence of robust justification. Mitigation strategies like PPI can be time-consuming, logistically complex, and yield diverse opinions. Proactively consideration of these complexities allows for setting realistic expectations and preventing surprises.

More impactful topics like conflict of interest and data governance—common in research conducted by commercial enterprises—may call for more careful consideration and reflections for their influence on review outcomes. Drawing from interviewee insights, we encourage digital health developers to adopt a more receptive stance towards information dissemination, seek external expertise, and engage with HEIs or other relevant entities for collaborative undertakings, as diverse team expertise facilitates tackling complex issues.

Satisfying ethical standards in research is a skill to develop. While ample guidance exists online, much of it is directed towards general research and remains in forms or on databases inaccessible to those outside the academic sphere ^43, 44^. Typically, industry professionals, like the interviewees, begin their search on RECs’ websites before turning to broader internet searches. Few possess the means or time to pursue literature research or entire books. The interviewees noted the absence of readily available guidance tailored to address specific queries unique to digital health.

Researchers have expressed the desire for more explicit guidance and open communication from ethics committees ^21^. The HRA has developed resources for the public, including some applicable to issues identified in the findings ^45^. Yet, the challenge arises when supporting a cohort in a domain with diverse study designs and ethical considerations not addressed by traditional guidance. A recurring request among interviewees is the provision of example submissions in digital health showing what good and bad looks like. While there may be reservations about releasing explicit examples, offering guiding principles supplemented with practical solutions can be beneficial. This could manifest as: “If your study encompasses X, consider Y. Prior applicants have navigated this by implementing Z.” Many interviewees emphasised the value of receiving feedback prior to formal submission. While preliminary feedback throughout the submission process would be beneficial, if resources are limited, clearly conveying to applicants that the review’s purpose is constructive rather than decisive to the final outcome could alleviate some anxiety.

Much of what has been highlighted so far is generalisable to all research. However, the digital health cohort in the UK faces unique challenges. Unlike some countries, the UK does not have independent IRBs that offer ethical review services for a fee ^46, 47^. While a few organisations or university departments occasionally make exceptions ^48–50^, these are not viable long-term solutions for digital health companies. Ethics governance in the industry has been described as the “wild west”. Large corporations may establish internal governance boards, but most digital health enterprises lack such capacity. If a study does not fall under the HRA’s purview, these enterprises face challenges in identifying alternative submission avenues. Some may forge affiliations with a HEI via collaborations, but others, especially those without academic backgrounds or networks, struggle. Calls in published literature advocate for fair access to ethical reviews, urging HEIs to broaden their view of ethics committees beyond just protective measures for their constituents ^46, 51^. The World Medical Association Declaration of Helsinki establishes the ethical imperative of timely public dissemination of findings ^52^. By sidelining enterprises from ethics reviews, a void is created, risking under-reviewed studies and potential risks to participants and hindering the publication of results. Though apprehensions exist regarding fee-based ethics review services, like perceptions of unfairness or contractual obligations ^53^, this research identifies an opportunity for RECs to weigh the merits of introducing paid services. The expressed readiness of enterprises to allocate funds for such services indicates a dual opportunity: enhanced access for digital health developers and a revenue stream to bolster REC services. It is worth noting that countries like the US have successfully adopted this model.

While enterprises may navigate their way to ethical reviews via the HRA or a HEI, their external status introduces hurdles in seeking approval. The language and tone of the REC can foster a sense of mistrust, amplifying the divide between researchers and RECs, when a collaborative relationship should be cultivated. There is a particular dilemma in discerning when an intervention benefits the majority versus when to uphold standards of equity and inclusion. Many RECs operate on a voluntary basis, and members bring a variety of experiences and perspectives. Social science research attests to the variation in individuals’ views stemming from their backgrounds, perceptions, and affiliations ^54, 55^,. Amid calls for more standardised procedures and reflections on REC functioning, this study does not venture into these intricate debates. However, it can spotlight barriers emerging from the distinct nature of technology.

The successful deployment and sustained operation of digital health interventions often require commercial backing. Most interventions found effective by research remain inaccessible to the general public ^56^. Numerous literatures highlighted the tech industry’s role in ensuring scalability and fostering sustainability through public-private partnerships ^57, 58^. Such collaborations are indispensable not only for advancing healthcare technology but also for ensuring sustainable implementation. A nuanced understanding of the practicalities underlying digital health development might foster a more accommodating stance on issues otherwise deemed untenable.

Historically, the tech industry has faced criticism for its data-handling practices. This is particularly challenging in healthcare, where sensitive issues and vulnerable populations necessitate utmost caution. Yet, as the convergence of digital technology and healthcare becomes inevitable, there is a need for an open-minded understanding of technological development requirements. The interviewees’ perspective offers a valid consideration: recognising the company’s motivations and where user protection aligns with its interests can cultivate a cautious, yet unbiased, approach.

Data management concerns did not carry heavy weight against favourable opinions, but were frequently mentioned. Recognising that much of digital health research is conducted by non-academic, non-NHS organisations, there is a need for greater adaptability towards data-handling practices unfamiliar to the HRA and HEIs. While reinforcing robust data governance remains central to participant safety, RECs are encouraged to approach without predisposed biases.

The rise in research projects with a primary digital focus necessitates updating practices. Practically, this calls for revamping resources, such as templates and forms, to encompass diverse study types, consent methodologies, and data storage methods; generating resources that directly address the subtleties of digital health interventions; and augmenting REC expertise in the digital health realm. Omitting irrelevant sections or offering optional fields in the application form may mitigate applicant frustrations. Further, distinguishing between various digital health interventions or delineating between digital health technology and medical devices, followed by bespoke guidance, could offer significant utility. Several interviewees voiced apprehensions regarding later amendments. Given digital health’s dynamic nature and frequent iterations, it is essential to streamline the amendment process. Initial measures may include enhancing the user-friendliness of IRAS. Beyond this, applicants would benefit from mechanisms that assess and permit pre-specified changes without needing amendments. This entails devising an effective process and providing clear guidance on the boundaries of permissibility.

Interviewees generally held an understanding attitude and acknowledge the importance of ethics reviews for ensuring safety, beneficence, and integrity. Those well-acquainted with research often describe the process as straightforward. However, many still reported hesitancy towards engagement, citing challenges and stressors encountered during the process. Beyond avoiding common pitfalls, a deeper understanding of the process could alleviate some concerns. A significant grievance voiced by interviewees is the prolonged wait for responses and the overall approval timeline. While addressing this directly might be challenging, more open communication about expected timelines could enable better planning and realistic expectations. Timeline guidance exists for other trial types ^59^, but digital health researchers might overlook them due to perceived irrelevance. As such, increasing transparency in the review process, detailing decision-making rationales, application approval rates, and the reasoning behind specific requests, could bolster confidence and counteract negative sentiments.

This was an exploratory study. We chose to focus on industry perspectives in the choice of interview participants. We were not able to interview REC participants. We mostly reviewed NRES documents and did not achieve the same degree of HEI REC representation.

Further examination into specific digital health subsets, such as artificial intelligence, is recommended to explore their unique intricacies. Future studies might also explore the amendment process. Many of our recommendations would need iterative development to ensure feasibility.

## Conclusion

We have highlighted the challenges in obtaining ethics approval in digital health research, some universal and others unique to commercial enterprises. There are opportunities for applicants to prepare for a smoother experience, such as grounding their study designs in established evidence and guidelines, and in PPI recommendations. Concurrently, UK RECs can enhance the process by offering adapted guidance, expanding access, and adopting a tailored, non-biased approach. In light of this, both parties are encouraged to foster a collaborative relationship through open communication, flexibility in perspectives, and a deepened mutual understanding of the ethical landscape in digital health. We plan to develop these findings into accessible guidance. There is an open avenue for implementing and evaluating the proposed changes’ effectiveness.

## Acknowledgements

We thank Paulina Bondaronek (UCL) and Hanjun Gu (UCL) for their assistance in the research process, and Matthew Darlison (UCL) for comments on an earlier draft. We thank the HRA, particularly Zoher Kapacee, Susannah Keeling, Ashley Totenhofer, Gemma Warren, Rebecca Evans, Jonathan Fennelly-Barnwell and Stephen Tebbutt. We also thank all the interviewees for their support and contributions to the data collection.

## Data availability

Interview data and data not obtained by freedom of information requests is not available for secondary use.

## Appendix– Semi-Structured Interview Protocol

### INTRODUCTION

Recap of the purpose of the study and the interview.

- Our study aims to understand the challenges of the ethics application process for digital health research and projects that had to apply for ethical approval.
- We are looking to analyse the ethics application documents to understand the issue objectively, then interview the applicants about your experience with applying for ethics through the interview.
- The terms research, project, study, evidence generation, will refer to the project for which you applied for ethics.
- Outline of the interview sections.
- Informed consent to record.

### INTERVIEW

#### Section 1: Background Information

1. Please can you confirm your role in X company
2. How big is the company? (Number of employees)
3. What was the location of the project you had to get ethics for?
4. When did the project take place?

#### Section 2: Study Overview

1. Briefly, tell me about your company and the digital health product or intervention the project was on. *Suggested follow-up questions:*
  a. Was/is the intervention/product available on the market? Is it available through the NHS (e.g. commissioned) or available directly to consumers (e.g. Available through app store)?
  b. Was the project sponsored? If so, was the sponsor of the study from a commercial, academic, or another non-commercial source?
2. What were the ethical considerations associated with the project? Were there any that were unique or specific to the digital health context?

#### Section 3: Experience of the Ethics Application (Including Barriers and Enablers)

1. Please describe your ethics application process, including events that occurred throughout the process. *Clarifying examples:* Filled out application, submitted, received feedback, altered procedure, received decision, etc. *Suggested follow-up questions:*
  a. Was the project approved to go ahead in the end?
  b. What was your expectation or understanding of the ethics approval process and **how did the actual process differ?** For example, you can also talk about how you felt about the process.
2. What do you think **went well** with the ethics application process? *Clarifying examples:* Please consider how you addressed the ethical issues, things you did during the application process, things the REC did that helped, and resources or help they provided, and anything else that stand out to you.
3. What were the **biggest challenges or barriers** you encountered during the ethics application process?

#### Section 4: Knowledge, Resources and Capabilities

1. What **internal expertise** did you have in your company surrounding ethics in digital health
2. What **internal resources** did you have for the ethics application and surrounding ethical considerations in your evaluation? (e.g. financial, human capital)
3. What **external support and resources** did you have or used throughout the process (e.g. links to universities or organisations, government frameworks, funding, etc.)

#### Section 5: Conclusion and Recommendations

1. In the context of digital health research/project/evidence generation, how would you like the ethics application process to change?
2. Is there anything that we haven’t talked about today on the ethics approval process in digital health that you would like to discuss?

### CONCLUSION

- Thank the participant for joining.
- Inform the participant that the final report will be circulated via email once produced.

## References

1. FDA. What is Digital Health? | FDA, https://www.fda.gov/medical-devices/digital-health-center-excellence/what-digital-health (2020, accessed 13 January 2023).

2. Meskó B, Drobni Z, Bényei É, et al. Digital health is a cultural transformation of traditional healthcare. Mhealth 2017; 3: 38–38.

3. Peek N, Sujan M, Scott P. Digital health and care in pandemic times: impact of COVID-19. BMJ Health Care Inform 2020; 27: 100166.

4. Murray E, Hekler EB, Andersson G, et al. Evaluating Digital Health Interventions: Key Questions and Approaches. American Journal of Preventive Medicine 2016; 51: 843–851.

5. Greaves F, Joshi I, Campbell M, et al. What is an appropriate level of evidence for a digital health intervention? The Lancet 2018; 392: 2665–2667.

6. Karpathakis K, Libow G, Potts HWW, et al. An evaluation service for digital public health interventions: User-centered design approach. J Med Internet Res; 23. Epub ahead of print 1 September 2021. DOI: 10.2196/28356.

7. Unsworth H, Dillon B, Collinson L, et al. The NICE Evidence Standards Framework for digital health and care technologies – Developing and maintaining an innovative evidence framework with global impact. Digit Health; 7. Epub ahead of print 2021. DOI: 10.1177/20552076211018617.

8. NHS England. The multi-agency advisory service (MAAS), https://transform.england.nhs.uk/ai-lab/ai-lab-programmes/regulating-the-ai-ecosystem/the-multi-agency-advice-service-maas/ (2022, accessed 18 January 2023).

9. Hedgecoe A. “A Form of Practical Machinery”: The Origins of Research Ethics Committees in the UK, 1967–1972. Med Hist 2009; 53: 331.

10. Gelling L. Role of the research ethics committee. Nurse Educ Today 1999; 19: 564–569.

11. the British Psychological Society. Code of Ethics and Conduct, https://www.bps.org.uk/guideline/code-ethics-and-conduct (2018, accessed 14 January 2023).

12. British Educational Research Association (BERA). Ethical Guidelines for Educational Research, https://www.bera.ac.uk/publication/ethical-guidelines-for-educational-research-2018 (2018, accessed 14 January 2023).

13. Newson AJ, Lipworth W. Why should ethics approval be required prior to publication of health promotion research? Health Promot J Austr 2015; 26: 170–175.

14. Graf C, Wager E, Bowman A, et al. Best Practice Guidelines on Publication Ethics: a Publisher’s Perspective. Int J Clin Pract Suppl 2007; 61: 1.

15. HRA. Integrated Research Application System, https://www.hra.nhs.uk/about-us/committees-and-services/integrated-research-application-system/ (2021, accessed 11 September 2023).

16. HRA. Research Ethics Committee – Standard Operating Procedures, https://www.hra.nhs.uk/about-us/committees-and-services/res-and-recs/research-ethics-committee-standard-operating-procedures/ (2022, accessed 11 September 2023).

17. McCormick JB, Sharp RR, Ottenberg AL, et al. The Establishment of Research Ethics Consultation Services (RECS): An Emerging Research Resource. Clin Transl Sci 2013; 6: 40.

18. Goodman KW. Ethics in Health Informatics. Yearb Med Inform 2020; 29: 26–31.

19. Petersen C, Subbian V. Special Section on Ethics in Health Informatics. Yearbook of medical informatics 2020; 29: 77–80.

20. Tusino S, Furfaro M. Rethinking the role of Research Ethics Committees in the light of Regulation (EU) No 536/2014 on clinical trials and the COVID-19 pandemic. British Journal of Clinical Pharmacology 2022; 88: 40–46.

21. Brown C, Spiro J, Quinton S. The role of research ethics committees: Friend or foe in educational research? An exploratory study. Br Educ Res J 2020; 46: 747–769.

22. Keith-Spiegel P, Koocher GP, Tabachnick B. What Scientists Want from Their Research Ethics Committee. Journal of Empirical Research on Human Research Ethics 2006; 1: 67– 81.

23. Mcareavey R, Muir J. Research ethics committees: Values and power in higher education. Int J Soc Res Methodol 2011; 14: 391–405.

24. Al-Durra M, Nolan RP, Seto E, et al. Nonpublication Rates and Characteristics of Registered Randomized Clinical Trials in Digital Health: Cross-Sectional Analysis. J Med Internet Res 2018;20(12):e11924 https://www.jmir.org/2018/12/e11924 2018; 20: e11924.

25. Micca P, Cruse CB, Shukla M. Health tech investment trends: How are investors positioning for the future of health? Insights into the quickly emerging health tech sector A report from the Deloitte Center for Health Solutions, https://blogs.deloitte.com/centerforhealthsolutions/. (2020).

26. Cox S, Solbakk JH, Bernabe RDLC. Research ethics committees and post-approval activities: a qualitative study on the perspectives of European research ethics committee representatives. Curr Med Res Opin. Epub ahead of print 2022. DOI: 10.1080/03007995.2022.2115773.

27. FDA. Clinical Decision Support Software Guidance for Industry and Food and Drug Administration Staff, https://www.fda.gov/regulatory-information/search-fda-guidance-documents/clinical-decision-support-software (2022, accessed 13 January 2023).

28. Almeida D, Shmarko K, Lomas E. The ethics of facial recognition technologies, surveillance, and accountability in an age of artificial intelligence: a comparative analysis of US, EU, and UK regulatory frameworks. AI and Ethics 2021 2:3 2021; 2: 377–387.

29. Cornock M. General Data Protection Regulation (GDPR) and implications for research. Maturitas 2018; 111: A1–A2.

30. ISO. ISO - ISO 27799:2016 - Health informatics — Information security management in health using ISO/IEC 27002, https://www.iso.org/standard/62777.html (2016, accessed 13 January 2023).

31. Jandoo T. WHO guidance for digital health: What it means for researchers. 10.1177/2055207619898984; 6. Epub ahead of print 8 January 2020. DOI: 10.1177/2055207619898984.

32. Marelli L, Lievevrouw E, studies IVH-P, et al. Fit for purpose? The GDPR and the governance of European digital health. Taylor & Francis 2020; 41: 447–467.

33. Lievevrouw E, Hoyweghen I van. The social implications of digital health technology, https://lirias.kuleuven.be/retrieve/547200 (2019, accessed 14 January 2023).

34. Samuel G, Derrick G. Defining ethical standards for the application of digital tools to population health research. Bull World Health Organ 2020; 98: 239–244.

35. Nelson WA. Necessary competencies for ethics committees. Skilled internal consultants vital to addressing ethics issues. Healthc Exec 2013; 28: 46–8.

36. Peute LW, Lichtner V, Baysari MT, et al. Challenges and Best Practices in Ethical Review of Human and Organizational Factors Studies in Health Technology: a Synthesis of Testimonies. Yearb Med Inform 2020; 29: 58–70.

37. Steinbrook R. Protecting research subjects—the crisis at Johns Hopkins. N Engl J Med 2002; 346: 716–720.

38. Wald DS. Bureaucracy of ethics applications. BMJ 2004; 329: 282–284.

39. Fiske A, Henningsen P, Buyx A. Your Robot Therapist Will See You Now: Ethical Implications of Embodied Artificial Intelligence in Psychiatry, Psychology, and Psychotherapy. J Med Internet Res 2019;21(5):e13216 https://www.jmir.org/2019/5/e13216 2019; 21: e13216.

40. Braun V, Clarke V. Using thematic analysis in psychology. Qual Res Psychol 2006; 3: 77– 101.

41. Fugard A, Henry PWW. Thematic Analysis. SAGE Research Methods Foundations: An Encyclopedia. Epub ahead of print 2019. DOI: 10.4135/9781526421036858333.

42. US FDA. Marketing Submission Recommendations for a Predetermined Change Control Plan for Artificial Intelligence/Machine Learning (AI/ML)-Enabled Device Software Functions. Draft Guidance for Industry and Food and Drug Administration Staff.

43. Greaney AM, Sheehy A, Heffernan C, et al. Research ethics application: A guide for the novice researcher. British Journal of Nursing 2012; 21: 38–43.

44. Remenyi D, Swan Nicola, Assem BVDen. Ethics protocols and research ethics committees : successfully obtaining approval for your academic research. 1st ed. Reading: Academic Publishing International Ltd., 2011.

45. HRA. Consent and Participant information sheet preparation guidance., https://www.hra-decisiontools.org.uk/consent/index.html (accessed 13 September 2023).

46. Brown N. Research ethics in a changing social sciences landscape. 10.1177/17470161221141011 2022; 19: 157–165.

47. Beckwith M. Errors in Trials. In: Operation Innovation: How to Make Society Richer, Healthier and Happier. London: The Entrepreneurs Network, 2023, pp. 32–34.

48. Independent researchers seeking ethics review. In: BPS Code of Human Research Ethics. British Psychological Society, 2021. Epub ahead of print April 2021. DOI: 10.53841/bpsrep.2021.inf180.12.

49. The University of Sheffield. Reviewing for external organisations. Research, Partnerships and Innovation.

50. Mitchels B. 7.1 Independent Review Panels (for research by independent practitioners). In: The Ethical Guidelines for Research in the Counselling Professions. Leicestershire: British Association for Counselling and Psychotherapy, 2019, pp. 73–74.

51. Young C. Could Ethics Committees please welcome Independent Researchers? Oral Health and Dentistry 2017; 1: 180–182.

52. World Medical Association. World Medical Association Declaration of Helsinki: ethical principles for medical research involving human subjects. JAMA 2013; 310: 2191–2194.

53. Pickworth E. Should local research ethics committees monitor research they have approved? J Med Ethics 2000; 26: 330.

54. Goffman E. Frame analysis: An essay on the organization of experience. Cambridge, MA, US: Harvard University Press, 1974.

55. Tajfel H, Turner JC. The Social Identity Theory of Intergroup Behavior. Polit Psychol 2004; 276–293.

56. Rogers MAM, Lemmen K, Kramer R, et al. Internet-Delivered Health Interventions That Work: Systematic Review of Meta-Analyses and Evaluation of Website Availability. J Med Internet Res; 19. Epub ahead of print 1 March 2017. DOI: 10.2196/JMIR.7111.

57. WHO. Global diffusion of eHealth. Report of the third global survey on eHealth 2016; 154p.

58. Free C, Phillips G, Watson L, et al. The Effectiveness of Mobile-Health Technologies to Improve Health Care Service Delivery Processes: A Systematic Review and Meta-Analysis. PLoS Med 2013; 10: e1001363.

59. HRA. Phase 1 clinical trials, https://www.hra.nhs.uk/planning-and-improving-research/policies-standards-legislation/phase-1-clinical-trials/ (2022, accessed 13 September 2023).

